# Premature adult mortality in India: What is the size of the matter?

**DOI:** 10.1101/2021.05.03.21256550

**Authors:** Chalapati Rao, Aashish Gupta, Mamta Gupta, Ajit Yadav

## Abstract

**Background:** Reducing adult mortality by 2030 is a key component of the United Nations Sustainable Development Goals (UNSDGs). Monitoring progress towards these goals requires timely and reliable information on deaths by age, sex, and cause. To estimate baseline measures for UNSDGs, this study aimed to use several different data sources to estimate subnational measures of premature adult mortality (between 30 and 70 years) for India in 2017.

**Methods:** Age-specific population and mortality data were accessed for India and its 21 larger states from the Civil Registration System and Sample Registration System for 2017, and the most recent National Family and Health Survey. Similar data on population and deaths were also procured from the Global Burden of Disease Study 2016 and the National Burden of Disease Estimates Study for 2017. Life table methods were used to estimate life expectancy and age-specific mortality at national and state level from each source. An additional set of life tables were estimated using an international two-parameter model life table system. Three indicators of premature adult mortality were derived by sex for each location and from each data source, for comparative analysis

**Results:** Marked variations in mortality estimates from different sources were noted for each state. Assuming the highest mortality level from all sources as the potentially true value, premature adult mortality was estimated to cause a national total of 2.6 million male and 1.8 million female deaths in 2017, with Bihar, Maharashtra, Tamil Nadu, Uttar Pradesh, and West Bengal accounting for half of these deaths. There was marked heterogeneity in risk of premature adult mortality, ranging from 351 per 1000 in Kerala to 558 per 1000 in Chhattisgarh among men, and from 198 per 1000 in Himachal Pradesh to 409 per 1000 in Assam among women.

**Conclusions:** Available data and estimates for mortality measurement in India are riddled with uncertainty. While the findings from this analysis may be useful for initial subnational health policy to address UNSDGs, more reliable empirical data is required for monitoring and evaluation. For this, strengthening death registration, improving methods for cause of death ascertainment, and establishment of robust mortality statistics programs are a priority.

**KEY MESSAGES:** *What is already known?:*

- Reliable measures of mortality at adult ages are required for evidence-based health policy, monitoring and evaluation of progress towards health-related UN SDGs.
- In the absence of reliable data from CRVS systems in many countries including India, these measures are largely derived from alternate data sources, data synthesis, or modelling methods.

*What are the new findings?:*

- This article presents a comparative analysis of measures of premature adult mortality from several data sources for India and its 21 larger states, examining their reliability and correspondence
- Following a conservative approach, the article proposes the maximum estimate of mortality between the ages of 30 and 70 years by sex for each location from any source as the potential baseline level of premature adult mortality around 2016-2017

*What do the new findings imply?:*

- Although each of the six data sources or estimation methods demonstrated some weaknesses, the adequate quality of data from the Civil Registration System (CRS) in several states suggests that through the implementation of strategic interventions, the CRS could be developed into a reliable data source for tracking progress towards the UNSDGs

## INTRODUCTION

Over the past three decades, there have been declines in child mortality across the world, as a result of global and local actions under the United Nations Millennium Development Goals campaign.(1, 2) Concomitantly, mortality at adult ages has been recognized as a growing component of population level disease burden.(3, 4) For this reason, the health-related United Nations Sustainable Development Goals (UNSDGs) for 2030 include specific targets to reduce adult mortality from major non-communicable diseases (NCDs), tuberculosis, HIV/AIDS, road traffic accidents, suicides, and occupational and environmental exposures, among other health conditions and risk factors.(5) Monitoring progress towards these UNSDG targets requires routine and reliable measures of population level cause-specific mortality at adult ages, for which Civil Registration and Vital Statistics (CRVS) systems with medical certification of cause of death are the optimal data source.(6) Unfortunately, reliable CRVS systems are not yet in place in many parts of the world, which limits the monitoring of adult mortality trends(7, 8)

Good quality CRVS data has been directly used to analyze national adult mortality trends in high income countries. (9) For several Latin American countries and South Africa, CRVS data were first adjusted for incomplete death registration, prior to similar trend analyses.(10, 11) The findings from these studies enabled an improved understanding of underlying diseases and risk factors for adult mortality in these countries. In India, although death registration systems have existed for over 150 years, not all deaths are currently registered in the national Civil Registration System (CRS).(12) Levels of death registration completeness vary by gender, age, and location across the country, and these data gaps have limited the reliability of direct adult mortality measures.(13)

The Sample Registration System (SRS) and National Family Health Survey (NFHS) programs were established in 1970 and 1992 respectively as alternate sources of mortality data, and had been specifically designed to enable reliable measurement of indicators of infant and under-five mortality.(14) Although the SRS and NFHS programs also compile information on adult deaths, the samples are not adequately powered for precise adult mortality measurement, especially at sub-national levels.(13, 14) Despite these limitations, there have been several attempts in the past decade to estimate national and subnational adult mortality rates using SRS, NFHS and CRS data, but with varying calculation methods, indicators, and adult age categories, which limits the comparability of results across methods or over time.(13, 15, 16) As a result, patterns and determinants of adult mortality are less well understood.

More recently, there have been two initiatives that employed data synthesis methods using various data sources to derive national and sub national estimates of population and deaths by age and sex. The state-level results from the “National Burden Estimates” (NBE) Study for 2017 were based on national level estimates of population and deaths derived for the United Nations World Population Project (WPP).(17, 18) Similar estimates of populations and deaths for each state are also available from the Indian State-level Burden of Disease initiative, which is a component of the Global Burden of Disease (GBD) Study for 2016, carried out by a collaboration between the Indian Council of Medical Research, the Public Health Foundation of India, and the Institute of Health Metrics and Evaluation, University of Washington, USA.(19, 20) The NBE and GBD synthetic estimates add to the available empirical data sources, to help understand levels of mortality at adult ages.

To understand and address biological, environmental, behavioral, socio-economic or health system factors that potentially influence patterns of disease burden and adult mortality across the country, there is an urgent need for reliable and timely gender specific subnational measures of adult mortality. The aim of this study was to conduct a comparative analysis of premature adult mortality from different data sources at national and state level around the period 2013-2017, to improve overall understanding of subnational levels and differentials in adult mortality patterns. We defined the interval between ages 30 and 70 years as the life span of premature adult mortality, and used three indicators to compare the magnitude of mortality at these ages across the different data sources and analytical approaches. This age interval corresponds to the indicator definition for NCD mortality under the UNSDGs.(21) We propose that our findings on the maximum plausible levels of total mortality from all causes in this age interval across all data sources could be used as a suitable proxy baseline measure for monitoring progress in India and its states towards the UNSDG NCD targets, till such time that detailed information on causes of death is available. The findings are also used to make recommendations for improving adult mortality monitoring levels in India, during and beyond the UNSDG era.

## METHODS

### Data sources

This analysis is based on life table derived summary mortality measures by sex for India and 21 large states, during the period 2013-2017. Table 1 presents the general characteristics of data on age-specific population and deaths from the CRS, SRS, NFHS, GBD, and NBE data sources.(13, 17, 19, 22, 23) that were used for life table analysis. Relevant additional details are also available from the GATHER Statement for this study (see Appendix 1). The SRS life tables were based on observed population and deaths in sample sites during 2017. For the NFHS life tables, the household survey data were used to create life-lines to estimate deaths and person-years lived in the exposure period. For the CRS life tables, recorded deaths in 2017 and population projections for 2017 developed by the National Commission on Population (NCP) were used.(24) The NBE and GBD studies reported estimates of deaths by age and sex for each state derived from the GBD and NBE internal modelling processes. (17, 19, 25). These studies also reported state specific populations by age and sex, derived from the United Nations World Population Prospects and the IHME international population models, respectively.(18, 20). The NBE data did not include separate estimates for states of Andhra Pradesh, Himachal Pradesh, Telangana, and Uttarakhand. We used these available GBD and NBE population and death estimates in our life table calculations to derive the GBD and NBE based life tables for each state. The SRS proportionate age distributions of deaths were used to interpolate the coarser age categories of deaths from CRS and NBE to the standard age categories for our life tables, which were as follows: 0 to 1 year, 1 to 4 years, 5 to 9 years ….80 to 84 years, 85+ years.

**Table 1:**
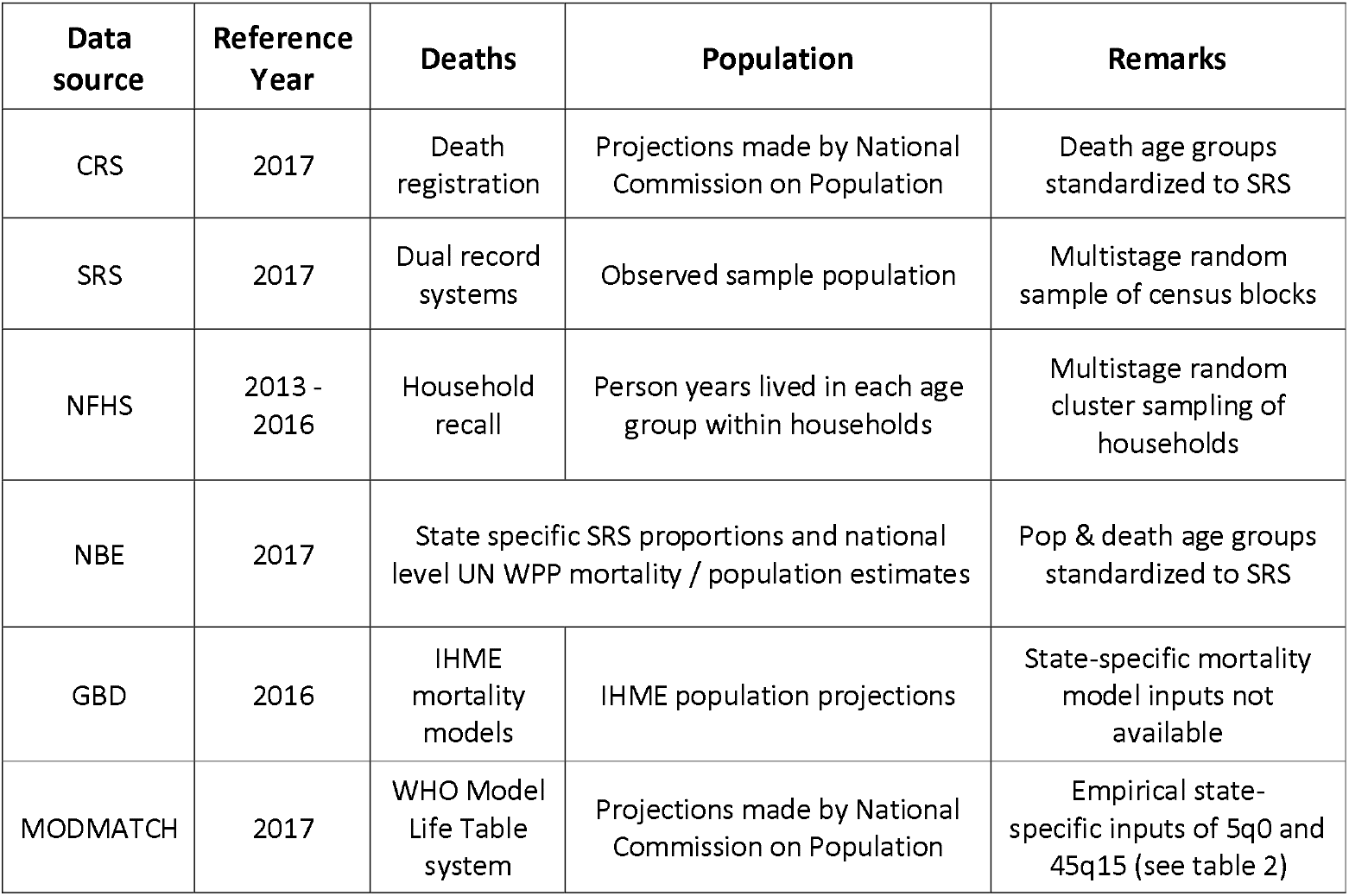
Characteristics of inputs used from different data sources to estimate life tables for India and states, 2013-2017

| Data source | Reference Year | Deaths | Population | Remarks |
| --- | --- | --- | --- | --- |
| CRS | 2017 | Death registration | Projections made by National Commission on Population | Death age groups standardized to SRS |
| SRS | 2017 | Dual record systems | Observed sample population | Multistage random sample of census blocks |
| NFHS | 2013 - 2016 | Household recall | Person years lived in each age group within households | Multistage random cluster sampling of households |
| NBE | 2017 | State specific SRS proportions and national level UN WPP mortality / population estimates |  | Pop & death age groups standardized to SRS |
| GBD | 2016 | IHME mortality models | IHME population projections | State-specific mortality model inputs not available |
| MODMATCH | 2017 | WHO Model Life Table system | Projections made by National Commission on Population | Empirical state-specific inputs of 5q0 and 45q15 (see table 2) |

### Life table analysis

Since life table quantities are not affected by the age structure of a population, period life tables are standard demographic tools which enable mortality comparisons across space and time. Period life expectancy at birth, for instance, is the average lifespan of a hypothetical cohort if they experienced the current set of age-specific mortality rates throughout their lifetime. Population and death data from each of the five sources for each location were used as inputs to construct life tables, using a standard spreadsheet program (26) From each data source-location-sex specific life table, the following summary mortality risks were extracted for comparison:

a. Life expectancy at birth (LE)
b. Risk of dying between birth and age 5 years (_5_*q*_*0*_)
c. Risk of dying between 15 and 60 years (_*45*_*q*_*15*_), conditional on survival up to age 15
d. Risk of dying between 30 and 70 years (_*40*_*q*_*30*_), (risk of premature adult mortality) conditional on survival up to age 30

Uncertainty intervals were estimated for each of the above parameters applying bootstrap methods using a publicly available programmed spreadsheet specifically designed for such analysis.(27) (see Appendix 2)

### Model life table estimation

In addition, we estimated a sixth set of life tables by sex for each location, using the World Health Organization Modified Logit Life Table System, implemented through its customized software tool named ‘MODMATCH’.(28, 29) This system uses input values of _5_*q*_0_ and _*45*_*q*_*15*_ in a statistical model, to predict a complete schedule of age-specific mortality risks for the location of interest. Mortality patterns in the MODMATCH model life tables approach are predicted from relationships between observed levels of child and adult mortality and their associated age-specific mortality patterns across all ages. For MODMATCH, the regression equations for these relationships were derived using a historical series of reliable life tables.(28) For the current analysis, selected input values of _5_q_0_ and _*45*_q_*15*_ for each state were chosen from the three empirical data sources (CRS, SRS and NFHS). We generally selected the higher value for each variable as available from the CRS and SRS, with a few exceptions. From each MODMATCH derived life table, the life expectancy at birth and risk of premature adult mortality were derived for comparative analysis. The same input parameters were also used in the two-dimensional Logarithmic Quadratic (Log-Quad) Model Life Table System developed by Wilmoth et al for a sensitivity analysis of MODMATCH outputs.(30) Given the little difference between these estimates, we utilized the MODMATCH results for further analysis. The estimated MODMATCH age-specific mortality risks were applied to respective age-specific populations from the NCP projections, to derive MODMATCH estimated numbers of deaths at each age.

### Data quality evaluation

The quality of mortality data from each source was evaluated by comparing the trajectory and slope of graphs of age-specific risks of dying between 30 and 70 years by sex, across all sources. The natural log of mortality rates from ages 30 to70 were plotted to evaluate if mortality rates from each source followed the Gompertz law of linear increase in log-mortality.(31, 32) Model plausibility, was also assessed by confirming that the age-specific trajectories of MODMATCH derived mortality rates for each state reflected their input values of _*45*_*q*_*15*_ from the CRS, SRS, or NFHS, respectively. The MODMATCH graphs were also compared with similar graphs by sex for each state that were derived from the Log-Quad Model, to evaluate model consistency.

### Premature adult mortality burden

The absolute numbers of observed or estimated deaths between 30 and 70 years in each state by sex from each source in 2017 were compared, as cross-sectional measures of premature adult mortality burden. In addition, the _*40*_*q*_*30*_ risks from each source were applied to a cohort of individuals aged 30 years in 2017, to estimate the potential numbers of deaths that would occur in this cohort over a forty-year period till 2057. The cohort comprised one-fifth of the state population aged 30 to 34 years in 2017 from the NCP projections, and the estimated deaths are based on the assumption that mortality levels will remain constant at the 2017 level over the ensuing four decades.

### Patients and public involvement

Patients or the public were not involved in the design, or conduct, or reporting, or dissemination plans of our research

## RESULTS

### Evaluation of model inputs

The plausibility of risks of _*5*_*q*_*0*_ (child mortality) and _*45*_*q*_*15*_ (adult mortality) from different sources were evaluated for each location, to select the most viable inputs for our model life table analysis. The general approach was to select the highest plausible value of each input parameter from the three empirical sources, to reflect the potential maximal levels of mortality for the study reference time period. Table 2 shows that the CRS risks of _*5*_*q*_*0*_ are implausibly low, due to known under-registration of infant deaths in all states(12, 13). The NFHS records higher levels of _*5*_*q*_*0*_ than the SRS in most states, except for males in Kerala, and for females in Andhra Pradesh, Gujarat, Haryana, Kerala, and Rajasthan. However, the NFHS rates relate to a period of 3 years prior to the survey (2013-2016), in addition to being subject to sampling error and recall bias. In comparison, the SRS has a much larger sample size, and is a continuous recording system with half yearly check surveys, therefore less subject to recall bias.(33, 34) For these reasons, the SRS _*5*_*q*_*0*_ values were chosen as inputs for MODMATCH in all states except Bihar, Jharkhand and Uttarakhand., for which the NFHS values were chosen, since the SRS values are implausibly low, as observed from other studies.(35) The modelled GBD and NBE _*5*_*q*_*0*_ risks demonstrate a relative difference > ± 5% when compared to the SRS values in most states. Since the NBE and GBD mortality risks are essentially outputs derived from other modelling processes, they were not considered as inputs for our model life table analysis.

**Table 2:** Estimated risks of dying before age five years and between ages 15 and 60 years from various sources for India and States during 2014-2017

| Location | Risk of dying between birth and five years |  |  |  |  |  |  |  |  |  | Risk of dying between 15 and 60 years |  |  |  |  |  |  |  |  |  |
| --- | --- | --- | --- | --- | --- | --- | --- | --- | --- | --- | --- | --- | --- | --- | --- | --- | --- | --- | --- | --- |
|  | Male |  |  |  |  | Female |  |  |  |  | Male |  |  |  |  | Female |  |  |  |  |
|  | CRS | SRS | NFHS | NBE | GBD | CRS | SRS | NFHS | NBE | GBD | CRS | SRS | NFHS | NBE | GBD | CRS | SRS | NFHS | NBE | GBD |
| India | 11 | **39** | 51 | 43 | 39 | 9 | **42** | 47 | 45 | 43 | 182 | **203** | 211 | 213 | 210 | 110 | **144** | 129 | 137 | 155 |
| Andhra Pradesh | 16 | **34** | 45 | NA | 29 | 13 | **29** | 27 | NA | 28 | **257** | 229 | 249 | NA | 205 | 153 | **153** | 134 | NA | 144 |
| Assam | 8 | **38** | 61 | 45 | 54 | 7 | **49** | 53 | 52 | 55 | 178 | **235** | 221 | 213 | 237 | 154 | **166** | 142 | 132 | 171 |
| Bihar | 5 | 34 | 61 | 42 | 47 | 5 | 40 | 53 | 43 | 55 | 107 | 160 | 225 | 151 | 151 | 69 | 137 | **174** | 132 | 152 |
| Chhattisgarh | 21 | **55** | 69 | 47 | 42 | 16 | **46** | 62 | 50 | 43 | 242 | **249** | 236 | 255 | 288 | 171 | **194** | 186 | 149 | 212 |
| Gujarat | 17 | **45** | 48 | 45 | 32 | 15 | **44** | 38 | 43 | 33 | 213 | **213** | 240 | 216 | 215 | 114 | **127** | 112 | 120 | 150 |
| Haryana | 4 | **37** | 35 | 42 | 32 | 4 | **45** | 41 | 44 | 38 | 196 | **237** | 216 | 242 | 227 | **132** | 130 | 119 | 132 | 143 |
| Himachal Pradesh | 11 | **31** | 43 | NA | 31 | 10 | **24** | 31 | NA | 28 | 165 | **167** | 220 | NA | 205 | 73 | **85** | 90 | NA | 96 |
| Jammu & Kashmir | 14 | **32** | 42 | 34 | 33 | 18 | **29** | 35 | 34 | 36 | 68 | **158** | 147 | 158 | 166 | 65 | **126** | 104 | 101 | 119 |
| Jharkhand | 9 | 32 | **51** | 38 | 35 | 8 | 38 | **56** | 42 | 37 | 138 | 170 | **190** | 178 | 168 | 83 | **154** | 128 | 146 | 182 |
| Karnataka | 16 | **31** | 36 | 33 | 29 | 13 | **30** | 29 | 37 | 27 | **249** | 212 | 193 | 241 | 246 | 132 | **142** | 96 | 150 | 181 |
| Kerala | 8 | **13** | 10 | 11 | 11 | 7 | **13** | 6 | 12 | 11 | **162** | 137 | 143 | 172 | 160 | 73 | **77** | 57 | 81 | 90 |
| Madhya Pradesh | 11 | **58** | 66 | 66 | 43 | 10 | **58** | 62 | 65 | 45 | 190 | **221** | 209 | 248 | 237 | 124 | **144** | 137 | 136 | 151 |
| Maharashtra | 11 | **19** | 32 | 21 | 24 | 10 | **21** | 21 | 22 | 24 | 164 | **170** | 183 | 193 | 190 | 106 | **121** | 91 | 127 | 140 |
| Odisha | 23 | **44** | 49 | 56 | 51 | 18 | **42** | 46 | 54 | 45 | **212** | 202 | 228 | 208 | 197 | 137 | **159** | 146 | 143 | 153 |
| Punjab | 6 | **24** | 29 | 26 | 30 | 6 | **27** | 37 | 31 | 34 | 198 | **241** | 193 | 208 | 203 | 134 | **142** | 118 | 125 | 143 |
| Rajasthan | 9 | **48** | 55 | 51 | 44 | 7 | **57** | 50 | 56 | 51 | **257** | 229 | 197 | 248 | 246 | 114 | **126** | 105 | 121 | 151 |
| Tamil Nadu | 12 | **23** | 34 | 21 | 15 | 11 | **22** | 24 | 21 | 15 | **239** | 201 | 209 | 206 | 218 | **127** | 111 | 117 | 119 | 143 |
| Telangana | 11 | **33** | 29 | NA | 24 | 12 | **34** | 40 | NA | 23 | 166 | **204** | 306 | NA | 184 | 124 | **135** | 148 | NA | 128 |
| Uttar Pradesh | 6 | **59** | 77 | 59 | 54 | 6 | **70** | 82 | 64 | 64 | 121 | **255** | 218 | 262 | 251 | 78 | **206** | 149 | 187 | 201 |
| Uttarakhand | 5 | 31 | **54** | NA | 28 | 5 | 26 | **49** | NA | 29 | 109 | **252** | 250 | NA | 237 | 52 | **155** | 115 | NA | 131 |
| West Bengal | 14 | **26** | 39 | 27 | 31 | 11 | **29** | 24 | 28 | 30 | 143 | **154** | 175 | 167 | 176 | 106 | **143** | 127 | 116 | 139 |

The CRS _*45*_*q*_*15*_ values are the highest of the three empirical sources for six states in males, and for two states in females, and were chosen as the most plausible inputs for MODMATCH for these states. For most of the other states, the SRS _*45*_*q*_*15*_ estimates were considered more plausible than those from the NFHS, due to its larger sample size, and for the compatibility of SRS measures with the time period of other data sources, for comparison. For Bihar and Jharkhand, the NFHS _*45*_*q*_*15*_ measures were deemed more plausible, given the recognized under-reporting of SRS deaths in these states.(35) Similar to the comparisons of under-five mortality risks, the GBD and NBE modelled values of _*45*_*q*_*15*_ differed from SRS values by > ± 5% in three-fourths of all states,

### Model evaluation

Figure 1 shows that MODMATCH derived mortality risks show a smooth exponential rise in mortality risks after age 40 years for Tamil Nadu, Punjab and Bihar, and correlate with respective _*45*_*q*_*15*_ inputs from the CRS, SRS and NFHS respectively. This is less obvious for Bihar, potentially due to the relative instability of NFHS mortality risk trends by age, as a result of low sample size. These graphs generally support the plausibility of MODMATCH derived age-specific trends as compared to the SRS, GBD and NBE trend lines, which do not appear to conform to the Gompertz law of smooth exponential increase in age-specific mortality (linear increase on the log-scale). Similar aberrations were observed in the graphs for all states (see Appendix 3). Using the same input parameters, the Log-Quad model outputs of life expectancy at birth and of age-specific mortality trends by sex for each state were very closely correlated to those from MODMATCH (see Appendix 4).

**Figure 1:**
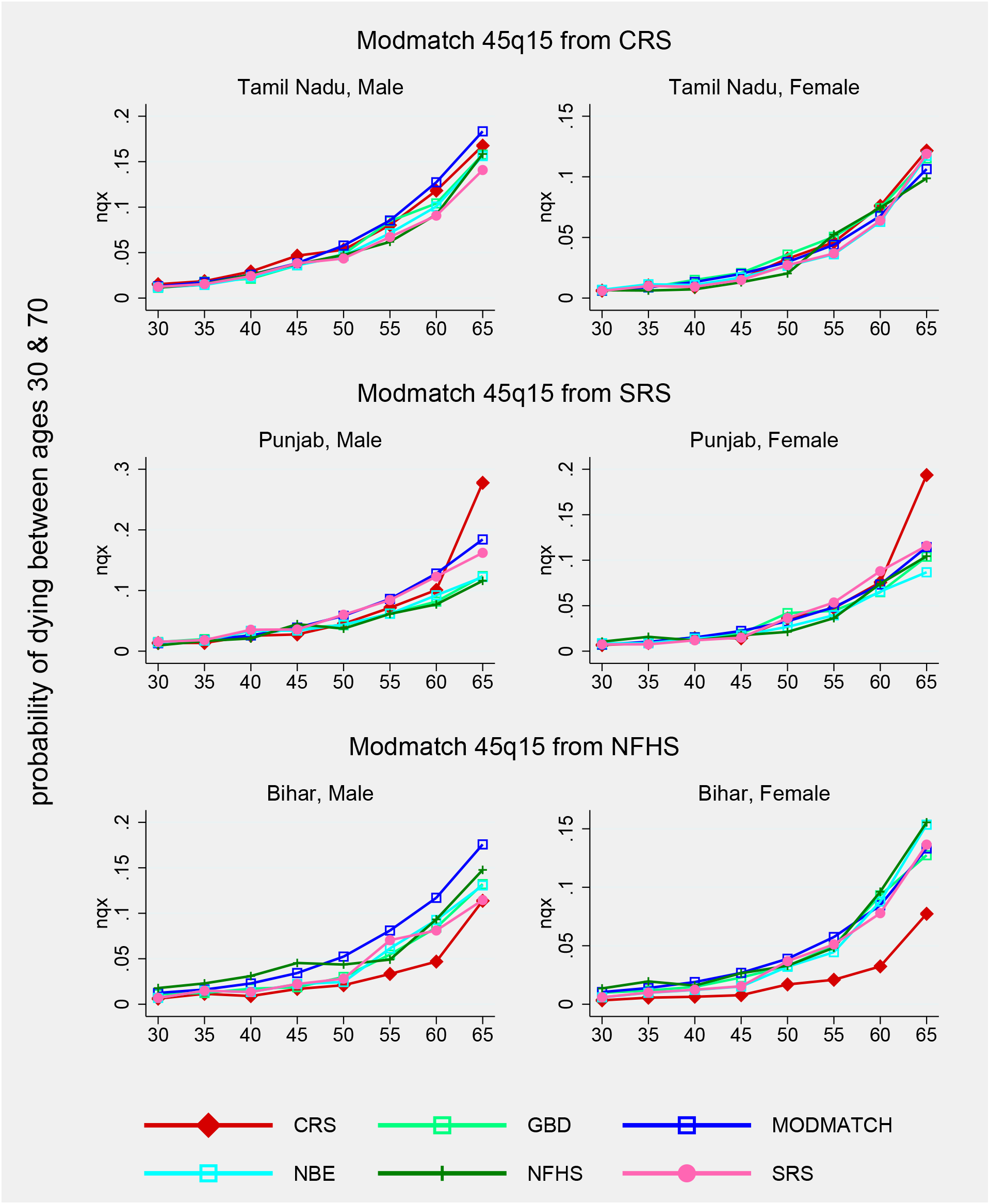
Mortality risk (30 to 70 years) from different sources for selected states, 2017

### Comparison of population exposures

National and state level differences in population exposures between the NCP projections, the NBE derivations from WPP estimates, and the GBD population estimates were evaluated to assess the likely impact of these denominators on the magnitude of deaths from each source. At the national level, the GBD population estimates are relatively higher than those from the NCP projections by 5.2%. Across states, these relative differences for males range from 1% in Kerala to 13% in Odisha, and in females from 2% in Kerala to 8% in Telangana. Similarly, the NBE populations also exceed the NCP projections by 2% at national level, with concomitantly lower orders of state level differentials, except for Jammu and Kashmir (12% for males, and 8% for females). These population differentials, although mostly in single digit percentages, actually translate into large population counts in most states, and strongly influence the comparisons of estimated deaths across data sources for each state.

### Overall mortality

CRS mortality reports were excluded from the comparisons of overall mortality, in view of the known under-registration of deaths in several states. Using the highest plausible empirically observed levels of _*5*_*q*_*0*_ and _*45*_*q*_*15*_ as model inputs, the MODMATCH life expectancies at birth were found to be lowest out of all the sources at national level. They were also the lowest in 17 states for males, and in 13 states for females. The MODMATCH age-specific mortality predictions are based on historical empirical mortality schedules from countries with complete mortality data, derived from modelled relationships between _*5*_*q*_*0*_, _*45*_*q*_*15*_ and overall mortality. Their plausibility is also supported by the findings from Figure 1 and Appendix 3, and suggest that life expectancies at birth could be potentially lower than what is known from the SRS, which is the national standard source for life expectancy estimates for India.(36) Even if the MODMATCH life expectancy estimates are not considered, there is a range of one year or more in life expectancies from the NBE, GBD and SRS for males in 13 states, and for females in 11 out of 21 states. These considerable differences in life expectancy at birth across the various sources of mortality estimates create uncertainty as to the true mortality levels for each state in 2017.

At the national level, MODMATCH estimated the highest number of total male deaths, while the GBD estimated the highest numbers of total female deaths. At state level, there were substantial differences in estimated deaths from each source, arising from variations in age-specific death rates as well as in estimated population exposures. The net effect of these variations was assessed in terms of the dispersion of estimated total deaths across sources. The dispersion was > 10,000 deaths in 18 out of 21 states for males, and 15 out of 21 states in females. Very high dispersions (>40,000 deaths) were observed for males in six states, and for females in four states. Such variations in total estimated deaths from different sources for each state also create uncertainty about the likely true levels of overall mortality at sub national level.

### Premature adult mortality risk

For interpreting the findings on premature adult mortality across different sources, a conservative approach was used in nominating the highest estimate for each state as the most likely value. Figure 2 shows considerable heterogeneity in the maximum values of _*40*_*q*_*30*_ mortality risks across the states. For males, these values range from 351 per 1000 in Kerala to 558 per 1000 in Chhattisgarh, and for females from 198 per 1000 in Himachal Pradesh to 409 per 1000 in Assam.

**Figure 2:**
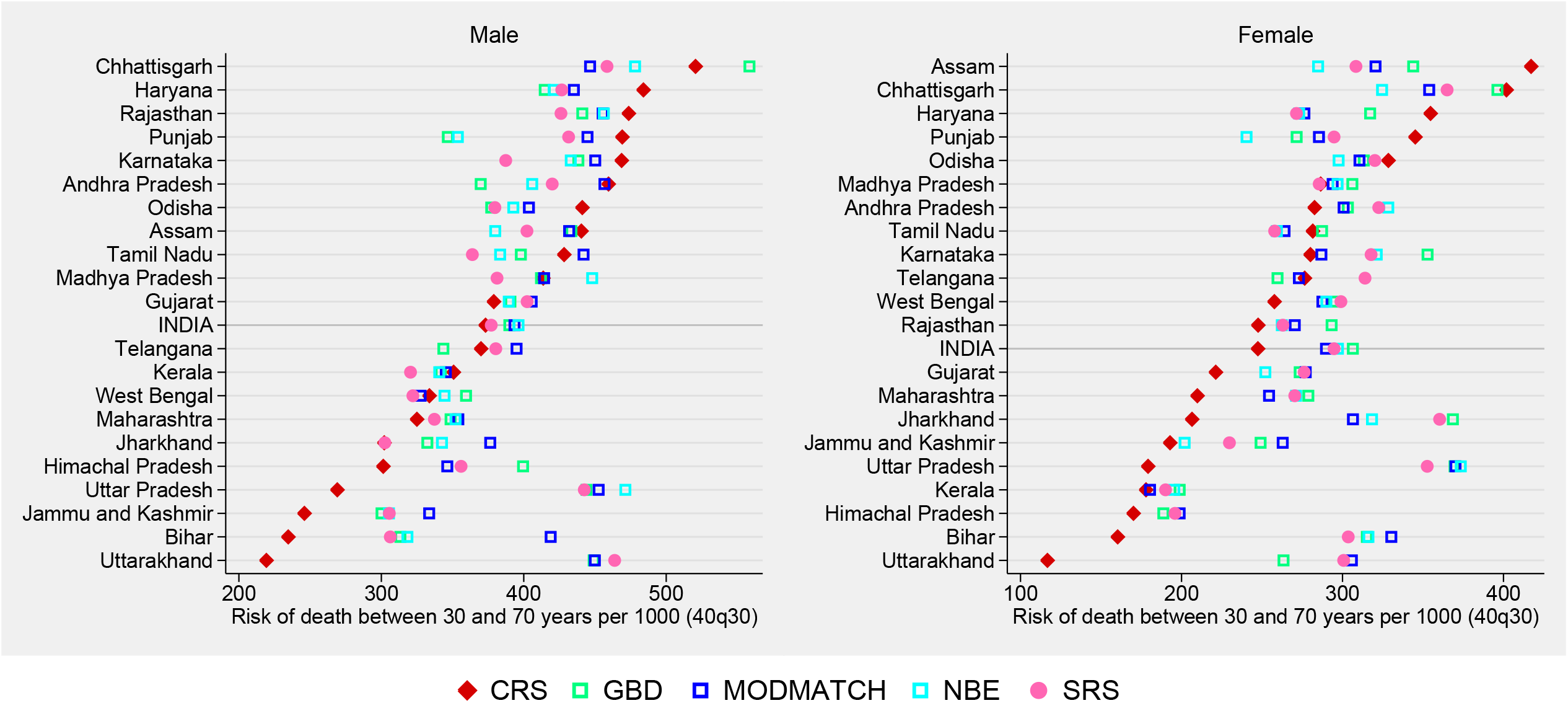
Estimated risk of dying between 30 and 70 years from different sources for India and states, 2017

On comparing estimated risks across data sources, Figure 2 shows that despite a general perception of incomplete death registration, CRS values of the _*40*_*q*_*30*_ mortality risks were the highest out of all sources for males in eight states (representing 28% of the national male population), and for females in five states (13% of national female population). The MODMATCH values were the highest values for males in eight states (42%), and for females in four states (11%). The GBD modelled _*40*_*q*_*30*_ estimates were the highest values for males in three states (10%), and females in seven states. (37%)

Another important comparison is between the CRS and SRS values. The observed CRS values of _*40*_*q*_30_ for males were higher than the SRS values in 13 states, and for females in seven states, indicating the likelihood of bias in SRS data, from sampling error and/or from problems with data quality. The graphs in Figure 2 show limited convergence in risks of premature adult mortality across data sources in several states for both males and females, even if the low CRS values for some states are not considered, due to incomplete death registration.

### Premature adult mortality burden

A cross-sectional perspective of the magnitude of premature adult mortality is provided in the left panel of Table 4, in terms of the maximum estimate of deaths between 30 and 70 years in each state during 2017, colour-coded to its data source. At the national level, a total of 2.6 million male and 1.8 million female deaths were expected to have occurred in this age group during 2017. Bihar, Maharashtra, Tamil Nadu, Uttar Pradesh and West Bengal account for approximately half of all these deaths. (See Appendix 5A for detailed estimates)

**Table 3:**
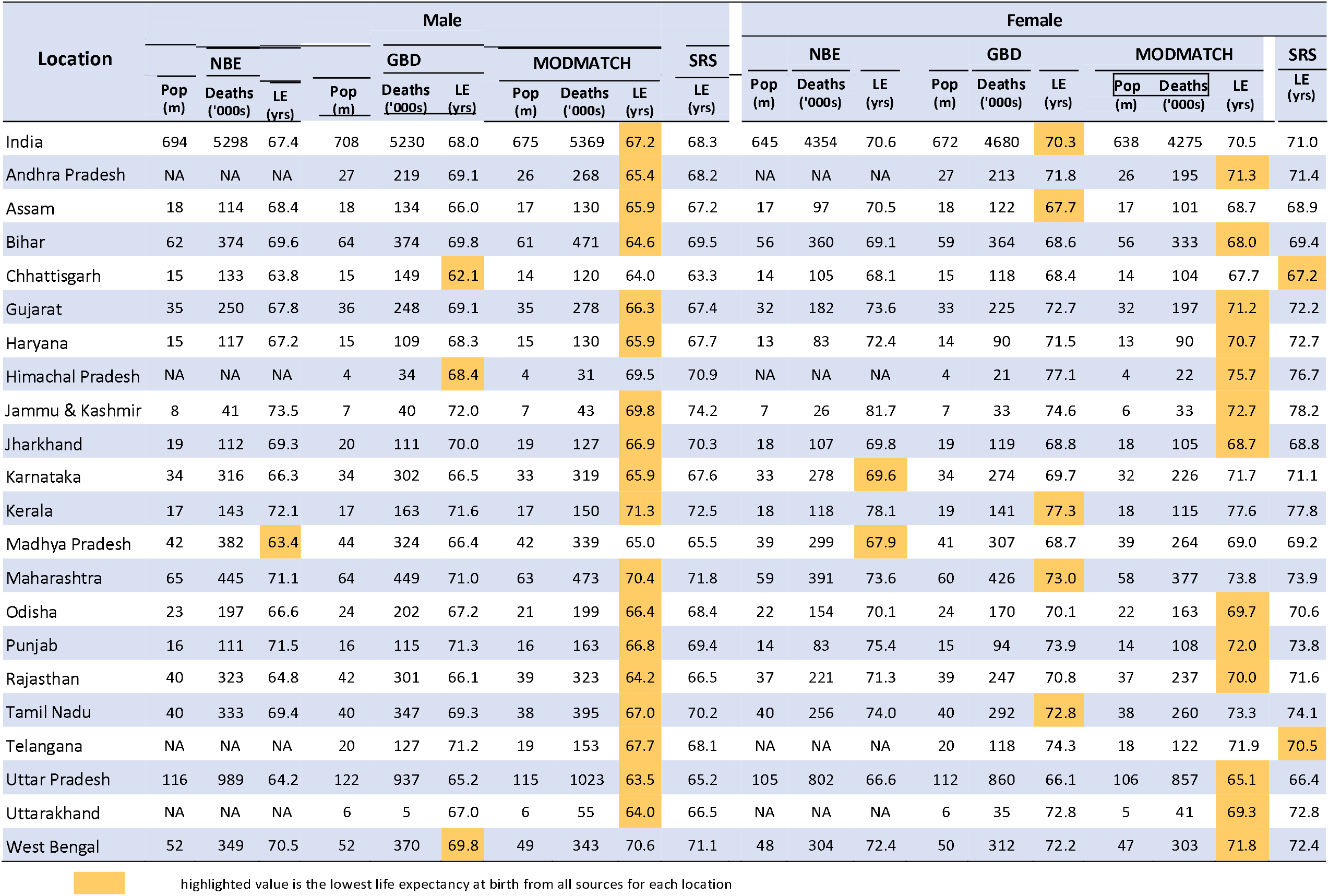
Estimated population, total deaths and life expectancy at birth by sex from various sources for India and States during 2014-2017

**Table 4:** Estimated deaths for India and states at ages 30-70 years in 2017, and expected cohort deaths during 2017 to 2057

| Location | Maximum estimates of premature adult mortality risks and deaths in 2017 |  |  |  | Expected number of deaths in a cohort of 30 year old men and women over the period 2017 - 2057 |  |  |  |
| --- | --- | --- | --- | --- | --- | --- | --- | --- |
|  | Men |  | Women |  | Men |  | Women |  |
|  | 40q30 per 1000 | Deaths ('000s) | 40q30 per 1000 | Deaths ('000s) | Cohort<sup>1</sup> ('000s) | Deaths ('000s) | Cohort<sup>1</sup> ('000s) | Deaths ('000s) |
| India | 396 | 2609 | 306 | 1802 | 10340 | 3859 - 4098 | 10045 | 2911 -3078* |
| Andhra | 459 | 135 | 323 | 85 | 434 | 160 - 199 | 448 | 126 - 135 |
| Assam | 440 | 66 | 417 | 50 | 269 | 102 - 118 | 277 | 79 - 116 |
| Bihar | 419 | 192 | 330 | 138 | 763 | 239 - 320* | 762 | 241 - 252* |
| Chhattisgarh | 558 | 79 | 402 | 53 | 212 | 95 - 119 | 209 | 68 - 84 |
| Gujarat | 406 | 130 | 276 | 83 | 572 | 217 - 232 | 515 | 130 -141* |
| Haryana | 484 | 63 | 355 | 39 | 243 | 101 - 117 | 217 | 59 - 77 |
| Hi machal | 400 | 16 | 199 | 7 | 59 | 18 - 24 | 60 | 10 -- 12 |
| Jammu and Kashmir | 334 | 17 | 263 | 12 | 110 | 27 - 37 | 98 | 19 - 26 |
| Jharkhand | 376 | 55 | 369 | 56 | 268 | 81 - 101 | 263 | 81 - 97* |
| Karnataka | 469 | 166 | 353 | 116 | 567 | 246 - 266 | 556 | 156 - 196 |
| Kerala | 351 | 74 | 199 | 43 | 242 | 82 - 85 | 276 | 49 - 55 |
| Madhya Pradesh | 448 | 164 | 306 | 99 | 614 | 253 - 275 | 575 | 165 - 176 |
| Maharashtra | 354 | 227 | 279 | 161 | 1071 | 348 - 379 | 979 | 249 - 273* |
| Odisha | 441 | 96 | 329 | 70 | 327 | 123 - 144 | 355 | 106 - 117 |
| Punjab | 469 | 71 | 345 | 46 | 257 | 91 - 121 | 240 | 58 - 83 |
| Rajasthan | 474 | 152 | 293 | 85 | 563 | 248 - 267 | 538 | 133 - 158 |
| Tamil Nadu | 442 | 197 | 287 | 126 | 624 | 248 - 276 | 652 | 169 - 187 |
| Telangana | 395 | 68 | 314 | 47 | 318 | 109 - 125 | 327 | 85 - 90 |
| Uttar Pradesh | 471 | 468 | 373 | 322 | 1540 | 684 - 726* | 1465 | 541 - 547* |
| Uttarakhand | 464 | 24 | 306 | 14 | 83 | 37 - 38* | 85 | 22 - 26* |
| West Bengal | 359 | 182 | 299 | 131 | 798 | 262 - 287 | 785 | 226 - 231* |
| <sup>1</sup> based on NCP population estimates |  |  |  |  |  |  |  |  |
| *CRS values not included due to low completeness of death registration |  |  |  |  |  |  |  |  |
|  | CRS | GBD | NBE | MODMATCH | SRS |  |  |  |

There is no single data source that consistently estimates the maximum number of deaths in all states. Although the GBD estimates the maximum deaths for females in two-thirds of all states, and for males in seven states, these are largely a result of the higher GBD population exposures. Similarly, NBE derived deaths are the highest at national level and five other states for males, also driven by higher population bases. On the other hand, the directly observed CRS deaths are the maximum for males in six states and for females in three states, and MODMATCH death estimates are maximum for males and females in three states each. The CRS and MODMATCH deaths are derived using the NCP predicted populations, which are the lowest of all three sources of population data.

The potential future burden from premature adult mortality till 2057 was calculated by applying the _*40*_*q*_*30*_ risks to a cohort of individuals from each state who were aged 30 years in 2017. On eliminating the differences from varying baseline population exposures, it was found that the maximum cohort estimates were mostly based on the MODATCH and CRS risks, particularly for males. (See Appendix 5B for detailed estimates). While these estimates of expected deaths offer some value in understanding the broad targets for mortality reductions in each state, the broad ranges of these estimates (approximately 20,000 deaths or more) across different sources for some states are a matter of concern, and will hamper the exact quantification of mortality reductions achieved, in the future, over the 40-year period till 2057.

## DISCUSSION

The principal findings from this analysis are the measures of premature adult mortality between ages 30 and 70 years for India and states from various data sources and methods around the commencement of the UNSDG era in 2017, as shown in Figure 2 and Table 4. Based on the assumption that the highest values of estimated mortality by sex for each state from any source represents this baseline measure for UNSDGs, a total of 2.6 million male and 1.8 million female deaths were expected to have occurred in India from this age group, during 2017. Bihar, Maharashtra, Tamil Nadu, Uttar Pradesh and West Bengal account for approximately half of all these deaths. In terms of mortality risks, there was considerable heterogeneity across states, with estimated values for males ranging from 351 per 1000 in Kerala to 558 per 1000 in Chhattisgarh, and for females from 198 per 1000 in Himachal Pradesh to 409 per 1000 in Assam.

Our findings show that at the national level, premature adult mortality accounts for nearly half of all deaths in males, and about a third of all female deaths, and these proportions are consistent across all the sources and for all states. (details in Appendix 5A). These large proportions actually translate into an annual burden of considerable numbers of deaths in each state each year, as shown in Table 4. These deaths are likely to be mostly from non-communicable diseases, injuries, tuberculosis and infectious hepatitis, which can be addressed through strengthening of ongoing programs for disease and injury prevention and control.(37-40) Also, our proposed estimates of deaths at these ages could be used as an outer bound of deaths to which cause-specific proportional mortality distributions from reliable epidemiological data sources could be fitted, to estimate the population level mortality burden from specific diseases and conditions.

Although the results have mostly focused on the comparisons and variations of estimates from different sources, our overall aim was to estimate the potentially true magnitude of premature adult mortality around 2016-2017. This is of critical importance, since these are required as baseline levels for monitoring the effectiveness of strategies to address health related UNSDGs that target this specific age category. Since each data source has limitations which vary in scope and effect across locations, it is not possible to nominate any single source as the optimal source for all states in 2017. Hence, we have chosen to report the maximum mortality estimate at these ages for each state, under the principle that health policies should be based on ‘worst case’ scenarios, for pragmatic health sector priority setting and resource allocation.

In this regard, we believe that the incorporation of mortality data from the Civil Registration System in 2017 in our estimation process has had an important influence on deriving our maximal mortality estimates. This is evidenced from the direct use of CRS derived values of _*45*_*q*_*15*_ risks as inputs for our model life tables for some states, as well as in applying the observed CRS _*40*_*q*_*30*_ risks in certain states to estimate the magnitude of premature adult deaths. Data from the Civil Registration System was not used for mortality estimation in the GBD and NBE analyses.

Three indicators were used to evaluate premature adult mortality, namely the risk of dying between the 30 and 70 years, the cross-sectional annual number of deaths at a baseline point in time, and a prediction of potential deaths in a cohort over a period of 40 years. Evaluating the risks of dying helps understand the underlying factors driving mortality, while cross-sectional estimates of deaths guide resource allocation for health services and clinical management to prevent death in the diseased, and predicted deaths serve as targets for disease control and mortality reduction strategies. Planning for disease prevention and control to reduce premature adult mortality is critical, since in addition to enabling individuals to fulfil their personal life goals, mortality prevention at these ages has important positive ramifications for their families, as well as society in general.

It is anticipated that the heterogeneity in mortality patterns is likely to be associated with varying disease profiles and causes of death patterns across the states. Hence, there would be a need for customized health programs for each state, and related indicators and targets to monitor and evaluate their impact. While the GBD and NBE studies have published state level epidemiological profiles, they differ in their respective cause-specific mortality patterns for individual states.(17, 19) Moreover, our analysis proves that even at a gross level, the GBD and NBE total estimated numbers of deaths for each state are not the same, and these are influenced by differing patterns of modelled age-specific mortality risks, and differing population exposures, resulting in disparate premature adult mortality risks. (see Appendix 6) These myriad explanations for differences in the GBD and NBE deaths for each state, and the absence of specific evidence to assess and justify the reliability of one source over the other, limits the utility of either of these sources, for establishing baseline levels of premature adult mortality to monitor progress towards the UNSDGs.

We have highlighted the lowest life expectancies at birth and the highest estimates of premature adult mortality by sex from all sources, as the most appropriate values to describe mortality levels for each state. The MODMATCH life expectancies at birth for most states are lower than what was previously understood from the SRS and GBD reports.(19, 36) However, the mortality measures from the CRS are known to be affected by bias due to under-registration. Our analyses did not include any measurements of registration completeness and therefore did not provide mortality estimates adjusted for such bias.

Our evaluation of plausibility of patterns of age-specific mortality risks (Figure 2) show that the GBD, NBE, and SRS age-specific mortality curves do not adhere to the Gompertz law that describes an exponential rise in mortality beyond 40 years, and such violations are generally considered to indicate problems with underlying data.(32) In contrast, the MODMATCH outputs of age-specific risks are compliant with the Gompertz law. Further, the results from MODMATCH were validated using the Log Quad Model Life Table System. We chose to use the MODMATCH results for our comparative analysis, since the details on state-specific inputs used in MODMATCH, and references to the software tool and relevant documentation are available in the public domain, to replicate and verify our results for each state.. Nevertheless, although the MODMATCH _*40*_*q*_*30*_ risks are generally higher than the SRS values for most states (see Figure 2), they are still below the same risks derived from observed deaths in the CRS, for both males and females in some states. These MODMATCH predicted mortality patterns are based on the expectation that current mortality in the study population reflects international historical trends, and this is not the case for these states with higher _*40*_*q*_*30*_ risks from empirical CRS data. Hence, the MODMATCH too potentially under-estimates premature adult mortality, and the degree of such under-estimation cannot be ascertained for several states with incomplete CRS death registration, particularly for females.

Another limitation of this analysis was our inability to establish the potentially minimum value of _*40*_*q*_*30*_ risk in Indian populations, due to all the uncertainty underlying our estimates. A reliable minimum risk could have been used as a counterfactual level, to estimate the potentially avoidable burden from premature adult mortality in each state.(41) For this estimation, the counterfactual minimum risk would be applied to each cohort of 30-year-olds in 2017, to yield the minimum expected deaths for each state over the 40-year period till 2057. Subtracting these minimum expected deaths from the cohort mortality estimates presented in Table 4 would yield the potentially avoidable burden from premature adult mortality in each state, if the state were to experience the same population health status and health system attributes as the population with the counterfactual risk. While it is possible to use an estimate of _*40*_*q*_*30*_ from a reliable international source as the counterfactual risk, we did not choose to do so, since it may not be epidemiologically coherent with Indian health experience.

From an overall perspective therefore, none of the available data sources or analytical methods appears to serve as the single appropriate source for understanding the true levels of premature adult mortality in India. Our approach to compare and report the maximum estimate for each state as the likely value is only borne out of expediency, to focus attention of health policy analysts and agencies on the potential magnitude of this component of disease burden around 2017, as baseline measures for the UNSDGs. In highlighting the gaps in empirical data, along with the variations in estimates across sources, we hope to draw the attention of public health bureaucrats to the urgent action required to strengthen the CRS as the optimal source for subnational mortality statistics in India. As described in detail elsewhere, the CRS is based on a sound legal framework and administrative structure, with adequate infrastructure and operations for basic registration of deaths by age and sex across the country, which have led to gradual improvements in data quality till date.(12, 13, 42) Further interventions are required to improve completeness of death registration in some locations, and attribution of causes of death more generally, based on an incremental sampling approach supported by adequate capacity building, over the next decade.(13, 43, 44) Also, CRS statistical reports should provide district level data, and with more detailed age-groups to directly estimate abridged life tables and other age related mortality measures. There is also a need to coordinate population projection exercises and methods across different groups, so that there is a unified set of consensus-based population exposures for evaluating mortality risks. The methods and findings from these analyses would be relevant for other countries facing the need to reconcile local mortality data with modelled mortality estimates from international sources.

## Data Availability

All primary data analyzed during this research is available in the public domain or can be obtained from data custodians whose details are made available within the reference list.

## CONTRIBUTORSHIP STATEMENT

CR conceptualized the research, led the analysis, and drafted the initial version of the manuscript. AG collaborated on the analysis and design of figures and tables and contributed to writing the manuscript. MG and AY provided inputs for the review of data quality, interpretation of results, and documentation of implications of the study findings in the discussion. All authors contributed to the preparation of the final version of the manuscript.

## ACKNOWLEDGEMENTS

None

