## Appendix 1 for "Premature adult mortality in India: What is the size of the matter?"

Appendix 1. GATHER checklist of information that should be included in reports of global health estimates, with description of compliance and location of information for article titled: Premature Adult Mortality in India: What is the size of the matter?

| **#** | **GATHER checklist item** | **Description of**  **compliance** | **Reference** |
| --- | --- | --- | --- |
| **Objectives and funding** | | | |
| 1 | Define the indicators, populations, and time periods for which estimates were made. | Information provided in Methods section | Methods section:  Populations and time periods for estimates from each data source described in first paragraph and Table 1  Indicator definitions   - summary life table measures: second paragraph - expected premature adult deaths: 7^th^ paragraph |
| 2 | List the funding sources for the work. | There were no funding sources | Not applicable |
| **Data Inputs** | | | |
| *For all data inputs from multiple sources that are synthesized as part of the study:* | | | |
| 3 | Describe how the data were identified and how the data were accessed. | Data on population and deaths obtained from  Government of India publications in public domain on Civil Registration System, Sample Registration System, National Family and Health Survey,  NBE data on population and deaths from website appendix of journal article mentioned as Reference 17  GBD data on population and deaths obtained through personal communication with authors of Reference 25 | Methods section lists references to data sources as cited in the text. Table 1 provides details of data sources |
| 4 | Specify the inclusion and exclusion criteria. Identify all ad-hoc exclusions. | All representative mortality data sources for India and states were included in this analysis, namely CRS, SRS, and NFHS. Synthesized mortality estimates from two additional studies (GBD, NBE) were also included for the comparative analyses. | Methods section. |
| 5 | Provide information on all included data sources and their main characteristics. For each data source used, report reference information. | Data sources and their characteristics are described in specific references for each source. | Characteristics of each data source relevant to this analysis are summarized in Introduction and Methods sections. Details of variables used from each source for analysis are provided in Table 1. Web links are included in the references for all data sources, for additional characteristics |
| 6 | Identify and describe any categories of input data that have potentially important biases (e.g., based on characteristics listed in item 5). | Summary of known biases in mortality data from empirical national data sources (CRS, SRS, NFHS)  included in methods.  For biases in mortality data from NBE and GBD, please see references 17 and 25 | The following potential data biases have been listed in the Introduction and Methods sections   a. Incomplete death registration in CRS b. Sampling error in SRS and NFHS c. Potential for recall bias in NFHS data |
| *For data inputs that contribute to the analysis but were not synthesized as part of the study:* | | | |
| 7 | Describe and give sources for any other data inputs. | This analysis did not use other data inputs that were not synthesized as part of the study | Not applicable |
| *For all data inputs:* | | | |
| 8 | Provide all data inputs in a file format from which data can be efficiently extracted (e.g., a spreadsheet as opposed to a PDF), including all relevant meta-data listed in item 5 | CRS, SRS and NFHS data inputs can be downloaded from the weblinks provided in the bibliography.  The NBE population and death estimates can be downloaded from the weblink provided to reference 17  Population and death inputs from the GBD for each state could be obtained by contacting the corresponding author of Reference 25. | Weblinks to data inputs available from references |

| **Data analysis** | | | | | | |
| --- | --- | --- | --- | --- | --- | --- |
| 9 | | | Provide a conceptual overview of the data analysis method. A diagram may be helpful. | The analysis involves computation of life tables using input data from various sources, and comparing the outputs. | | Conceptual overview of data analysis described in Methods section |
| 10 | | | Provide a detailed description of all steps of the analysis, including mathematical formulae. This description should cover, as relevant, data cleaning, data pre-processing, data adjustments and weighting of data sources, and  mathematical or statistical model(s). | Life table analysis follows standard commonly applied statistical methods. We used a standard spreadsheet tool for this analysis, which has programmed mathematical formulae for each step. We have provided a weblink to the spreadsheet tool, which also includes references to the background scientific documents for the methods.  We directly used secondary data as available from the provided references. The only element of pre-processing was the interpolation of CRS and NBE death count data from broad age groups into comparable finer age groups available from the SRS and GBD data sources, for comparable inputs of age-specific death rates for life table analyses. This interpolation step and the final age groups for analysis are described in the Methods section. | | Spreadsheet tool provided in reference 26  Interpolation step described in first paragraph of Methods section |
| 11 | | | Describe how candidate models were evaluated and how the final model(s) were selected. | We used the 2 parameter WHO Modmatch life table system to estimate abridged schedules of age-specific mortality by sex, for India and 22 states.  We used the same inputs parameters of 5q0 and 45q15 (numbers in bold font in Table 1) in the Log-Quad Model Life Table System (similar to Modmatch in regard to its function to predict abridged life tables for study populations).  Findings between the two sets of life tables were compared in terms of differences in Life expectancy at birth, and risk of dying between 30 and 70 years.  The findings were closely aligned with the results from Modmatch (see next point). We selected the Modmatch results for our comparative analysis, since all details of the Modmatch methods, software, and it’s application are available in the public domain, and we have provided weblinks to these materials. | | Details provided in Methods section. Weblinks to Modmatch materials are provided in the references.  Weblink to Log Quad model life table article provided in list of references. |
| 12 | | | Provide the results of an evaluation of model performance, if done, as well as the results of any relevant sensitivity  analysis. | Results of differences in life expectancy at birth by sex for all states Between Modmatch and Log Quad are provided in Appendix table 4A. We also compared the trends in age-specific mortality rates between 30 and 70 years from MODMATCH and Log Quad for each state, and found very close correspondence between them. Graphs available in Appendix 4B | | Appendix table 4A  Appendix Figure 4B |
| 13 | | | Describe methods for calculating uncertainty of the estimates. State which sources of uncertainty were, and were  not, accounted for in the uncertainty analysis. | Confidence intervals for summary life table outputs from each data source were calculated using a bootstrap method, following steps outlined in Andreev and Shkolnikov 2010 (Reference 27). We assumed uniform population distribution for all sources. | | Confidence intervals for all estimates presented in Appendix 2 |
| 14 | | | State how analytic or statistical source code used to generate estimates can be accessed. | Code available on request to corresponding author | | Methods |
| **Results and Discussion** | | | | | | |
| 15 | | | Provide published estimates in a file format from which data can be efficiently extracted. | Results are available in the main tables and appendix tables. | | Table 2-4, Appendices 2 and 5 |
| 16 | | | Report a quantitative measure of the uncertainty of the estimates (e.g. uncertainty intervals). | 95% confidence intervals of summary life tables measures provided in Appendix 2ta | | Appendix 2 |
|  | 17 | Interpret results in light of existing evidence. If updating a previous set of estimates, describe the reasons for changes in estimates. | | Interpretations from direct and comparative analysis of relevant mortality outcome indicators have been described in Results and Discussion sections. | Results and Discussion sections | |
|  | 18 | Discuss limitations of the estimates. Include a discussion of any modelling assumptions or data limitations that affect  interpretation of the estimates. | | Study limitations described in Discussion section | Discussion section | |
