## Appendix 4 for "Premature adult mortality in India: What is the size of the matter?"

**Appendix 4A:** Correspondence between Log-Quad and Mod-Match estimates for life expectancy at birth

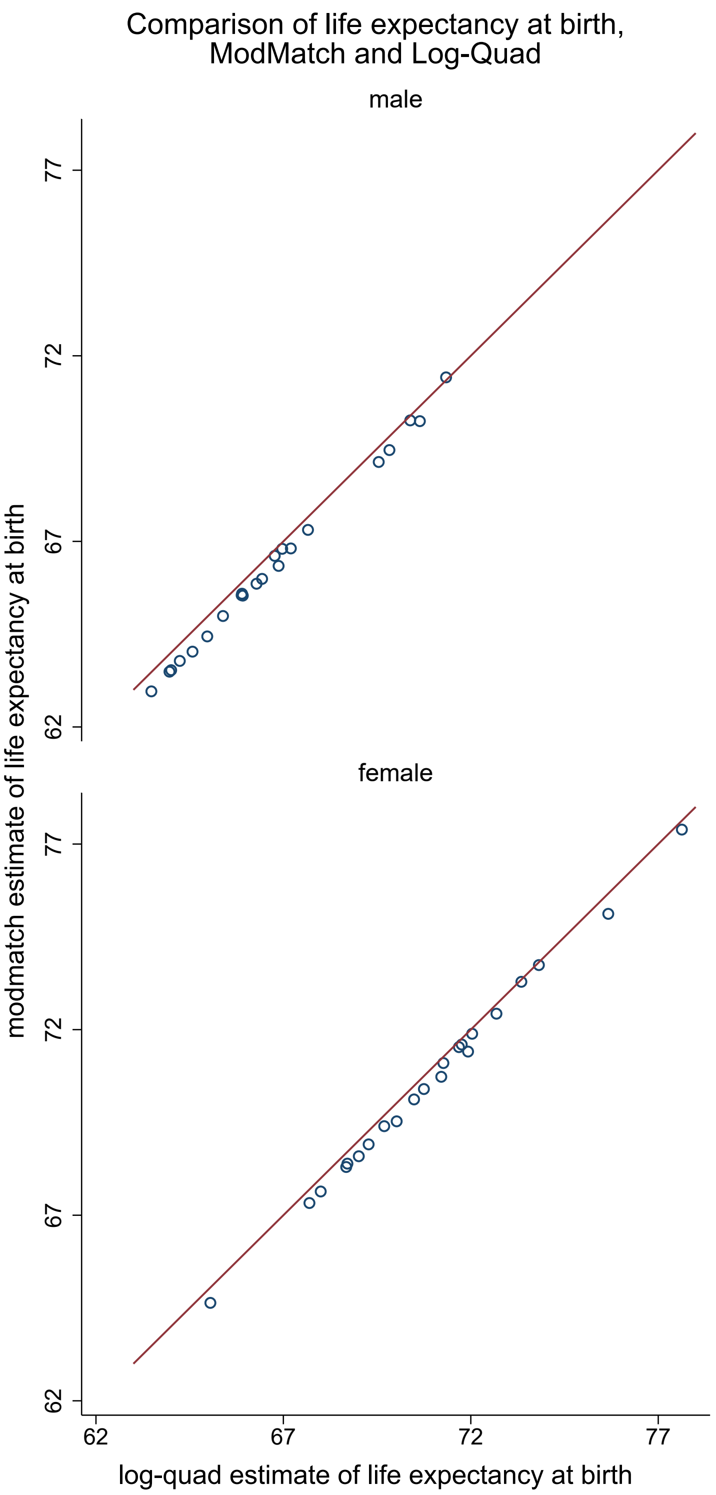

**Appendix 4B:** Correspondence between Log-Quad and Mod-Match estimates

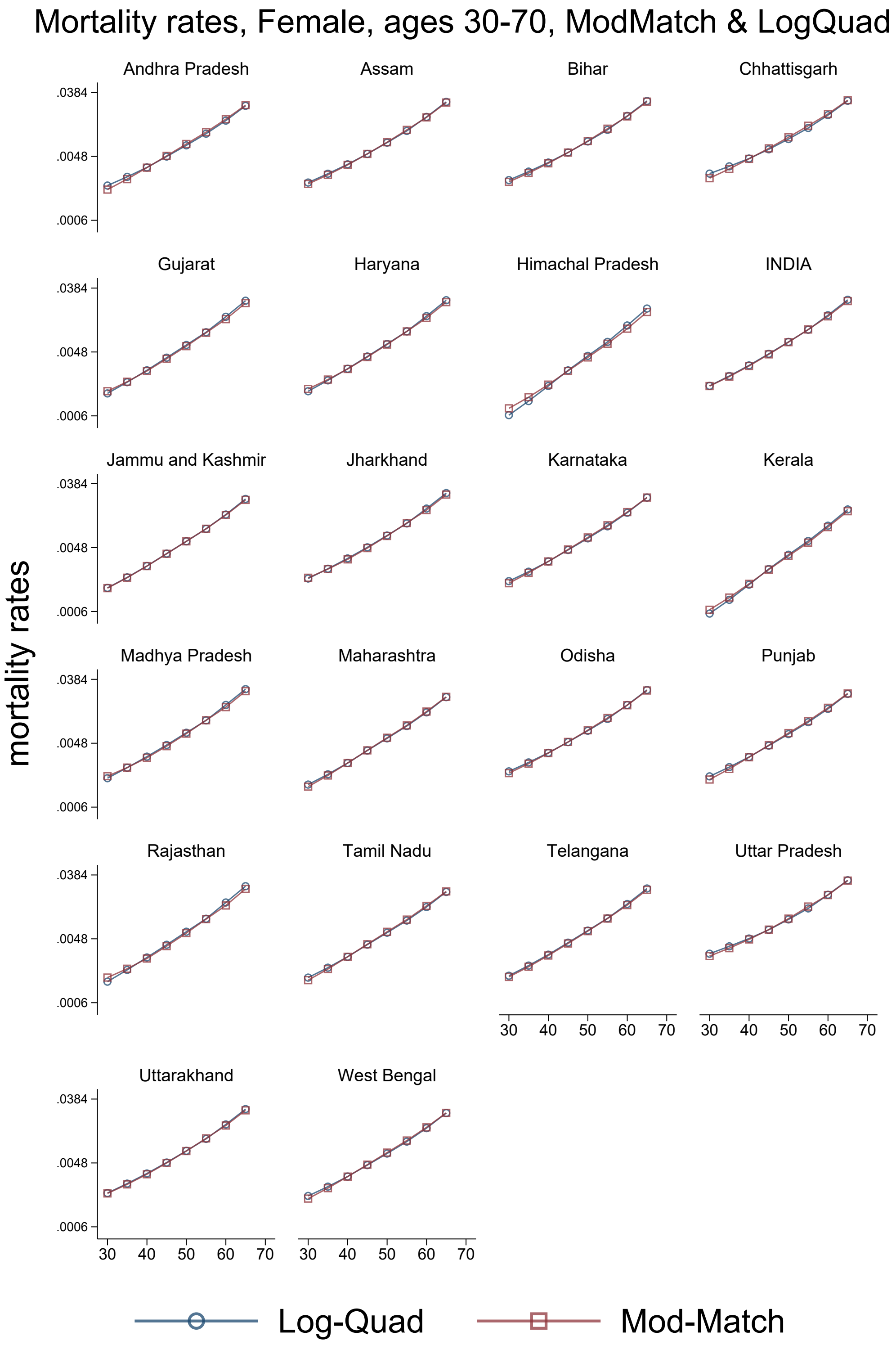

**Appendix 4C:** Correspondence between Log-Quad and Mod-Match estimates

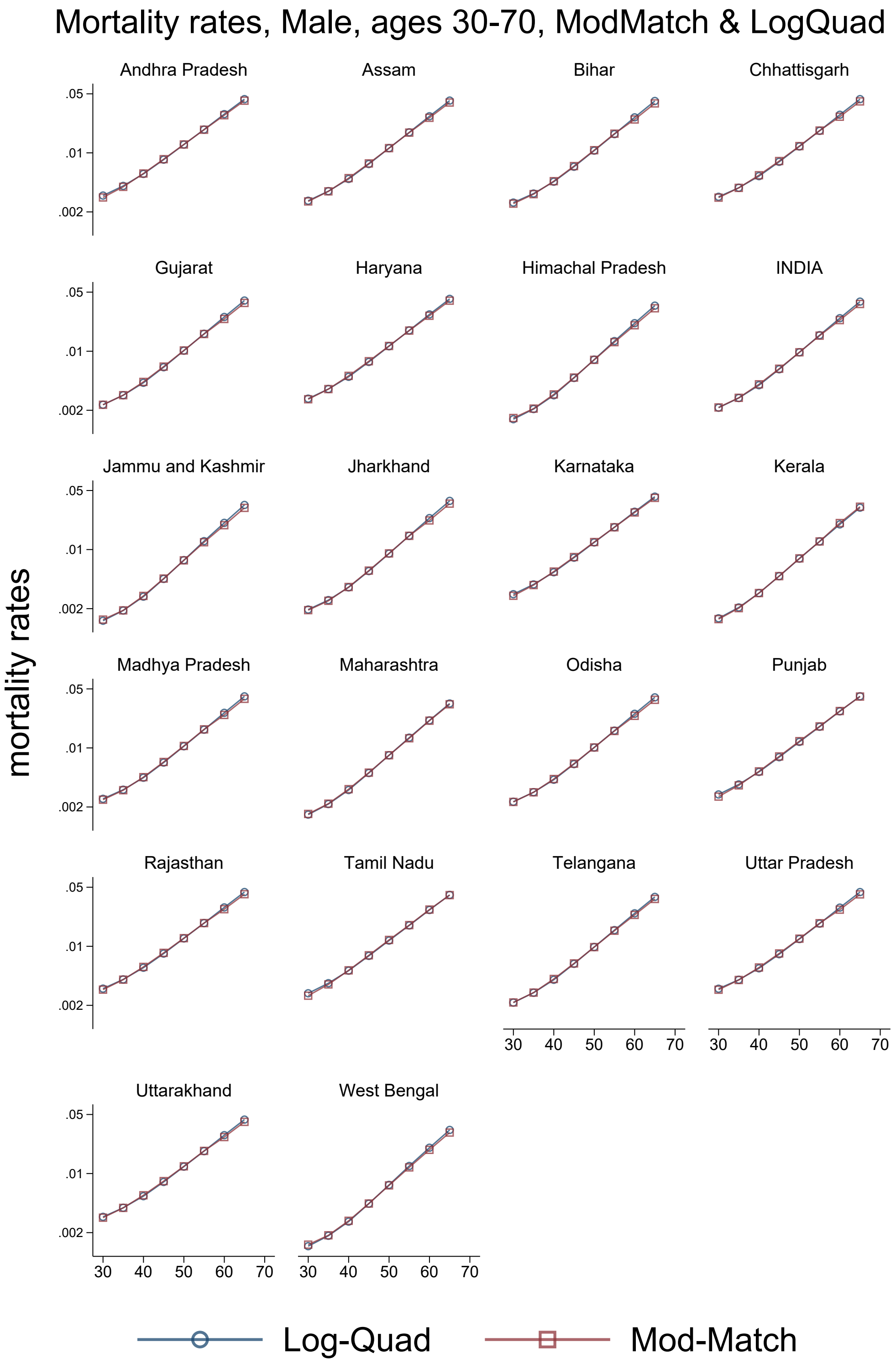
