## Supplementary figures and images for "Premature adult mortality in India: What is the size of the matter?"

### Appendix 6

**Appendix 6A:** Divergence between NBE and GBD estimates

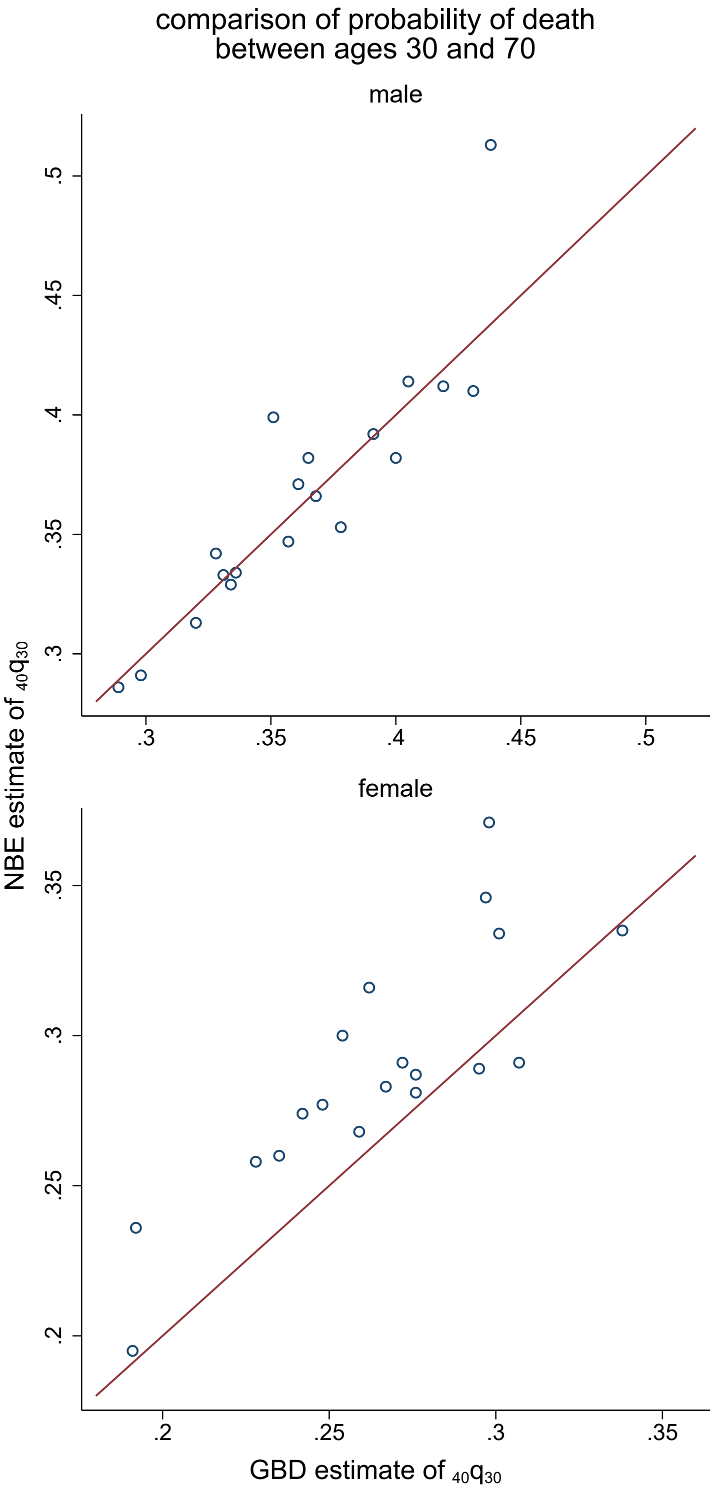
