## Appendix 3 for "Premature adult mortality in India: What is the size of the matter?"

**Appendix 3:** List of abbreviations, and age-specific mortality probabilities between ages 30 and 70, by source, sex, and state.

**List of abbreviations**

- 5q0: Probability of death between ages 0 and 5
- 45q15: Probability of death between ages 15 and 60
- 40q30: Probability of death between ages 30 and 70
- CRS: Civil Registration System
- CRVS: Civil Registration and Vital Statistics
- GBD: Global Burden of Disease
- IHME: Institute of Health Metrics and Evaluation
- LE: Life expectancy
- Log-Quad: Log-Quadratic Mortality Model
- MODMATCH: Modified Logit Life Table System
- NBE: National Burden of Disease Estimates
- NCDs: Non-Communicable Diseases
- NCP: National Commission on Population
- NFHS: National Family Health Survey
- SRS: Sample Registration System
- SVD-Comp: Singular Value Decomposition Mortality Model
- UNSDGs: United Nations Sustainable Development Goals

**Appendix 3A:** Mortality probabilities by place, source, and sex

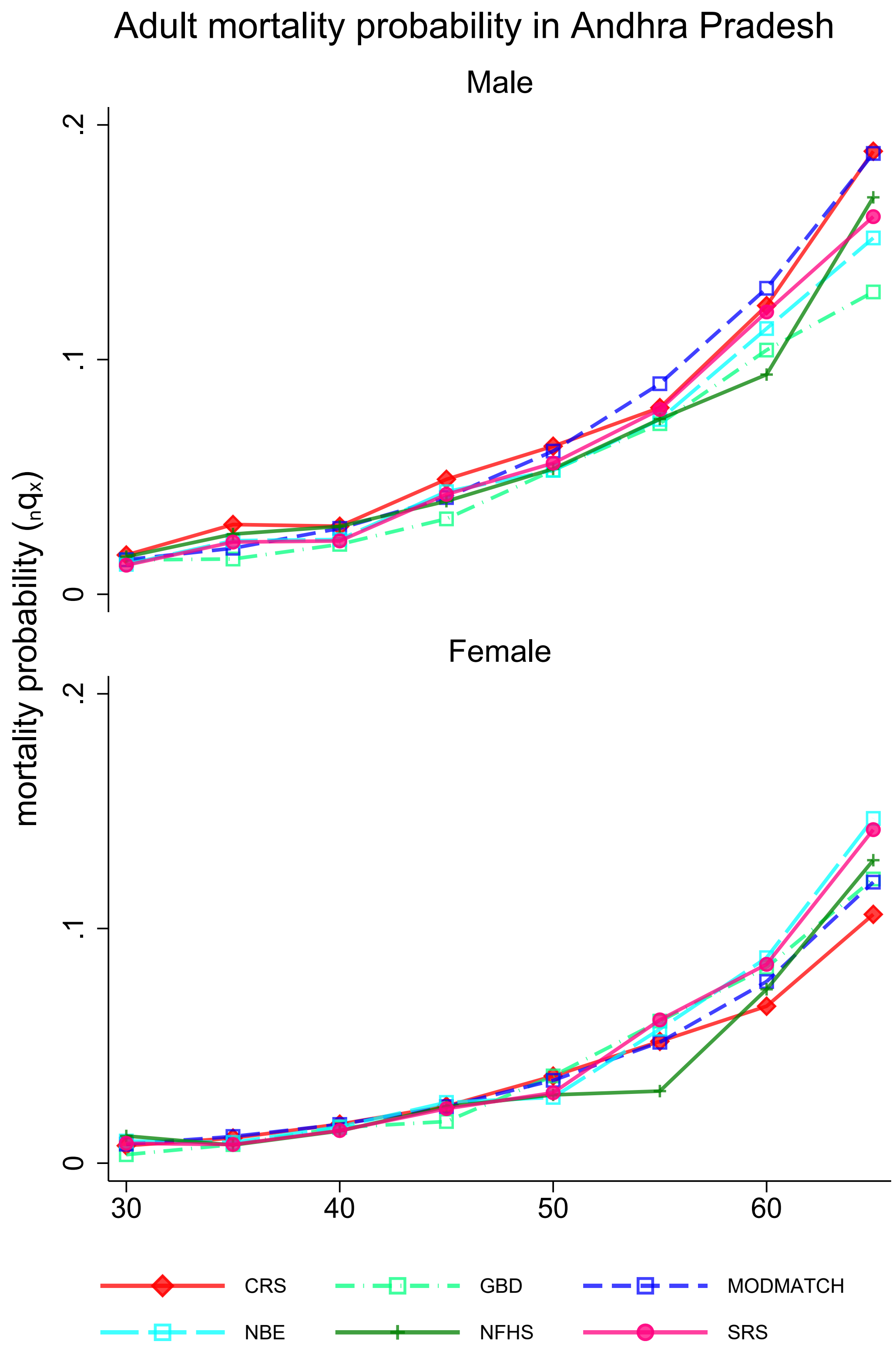

**Appendix 3B:** Mortality probabilities by place, source, and sex

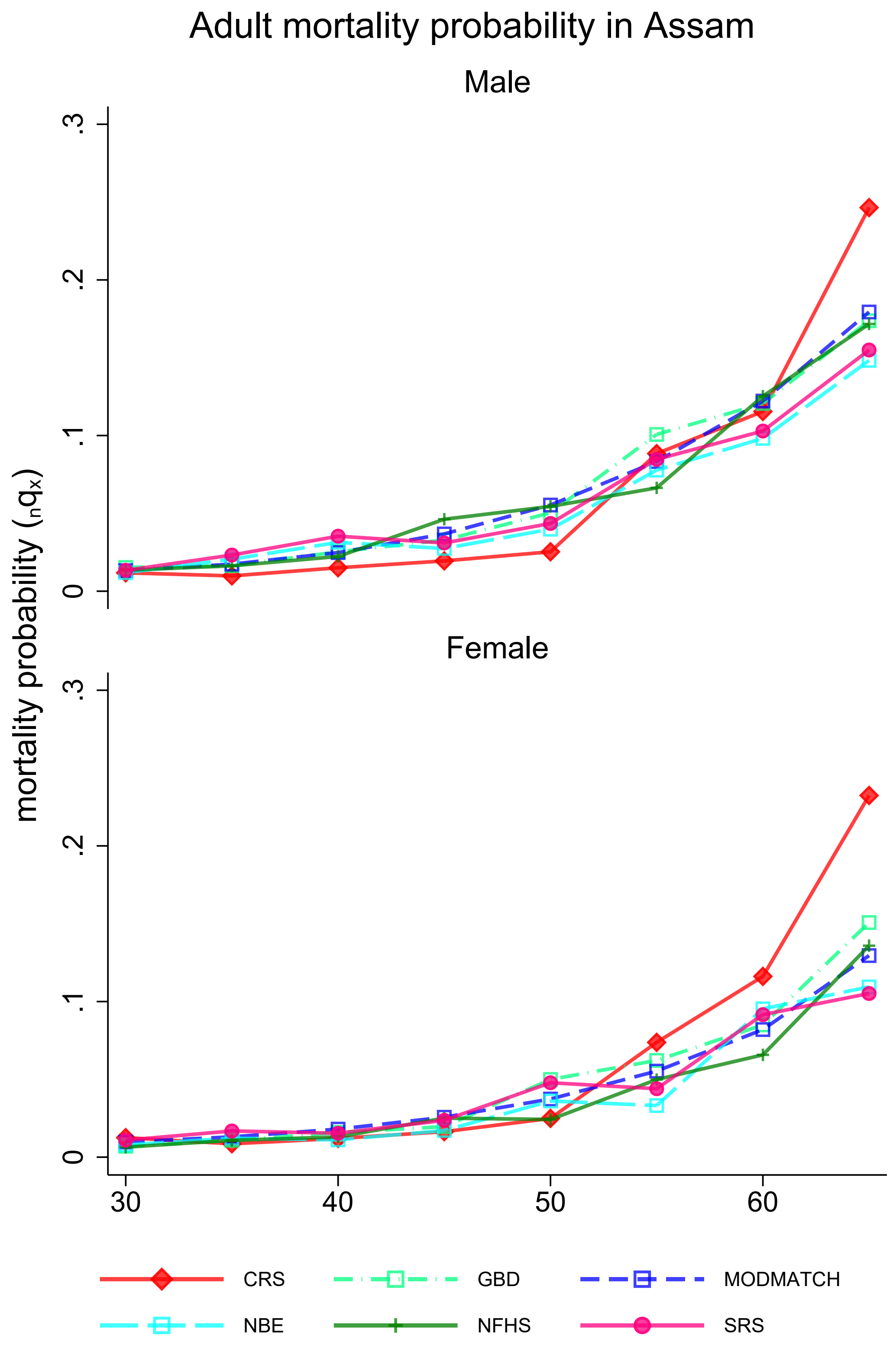

**Appendix 3C:** Mortality rates by place, source, and sex

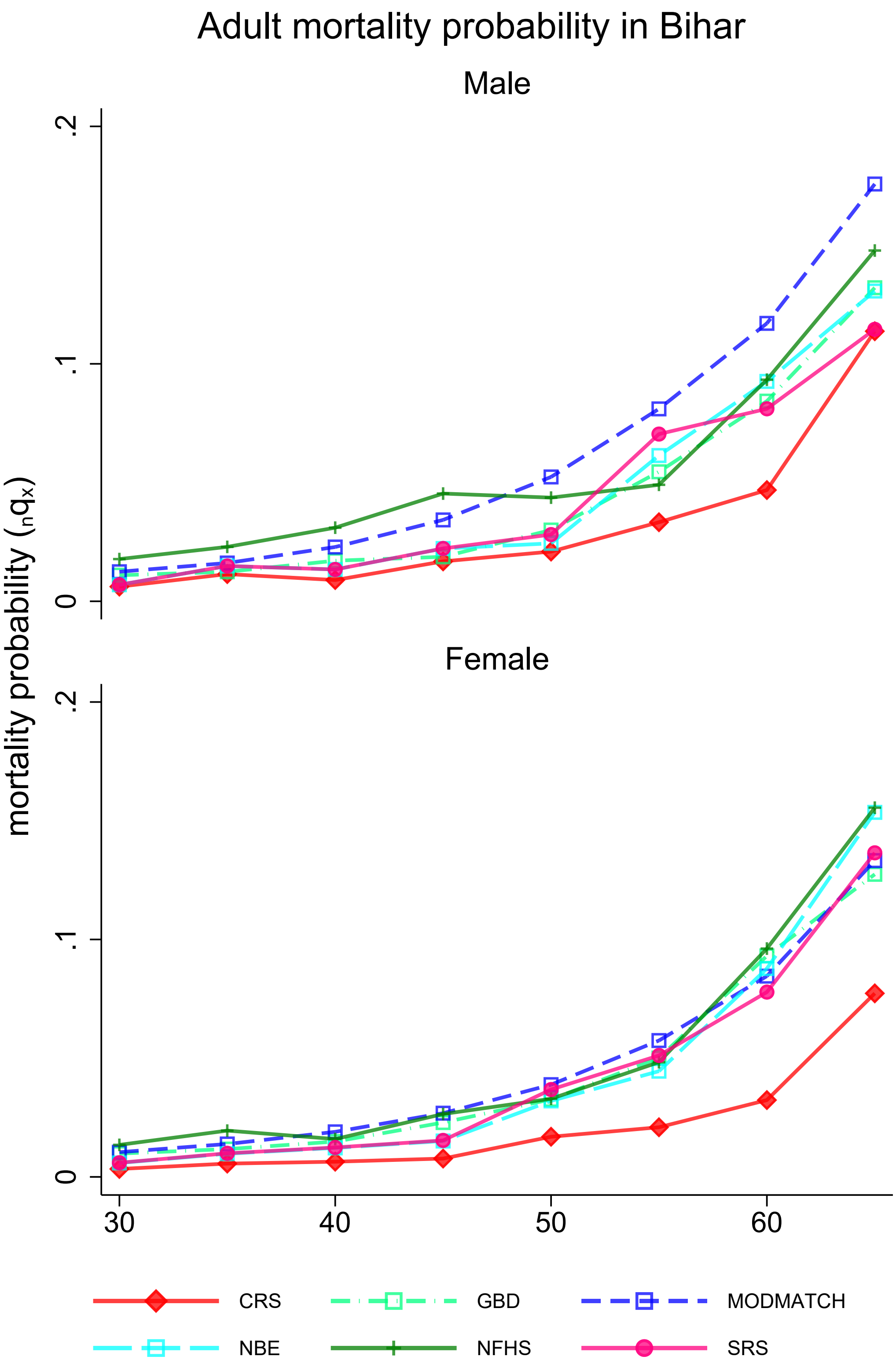

**Appendix 3D:** Mortality probabilities by place, source, and sex

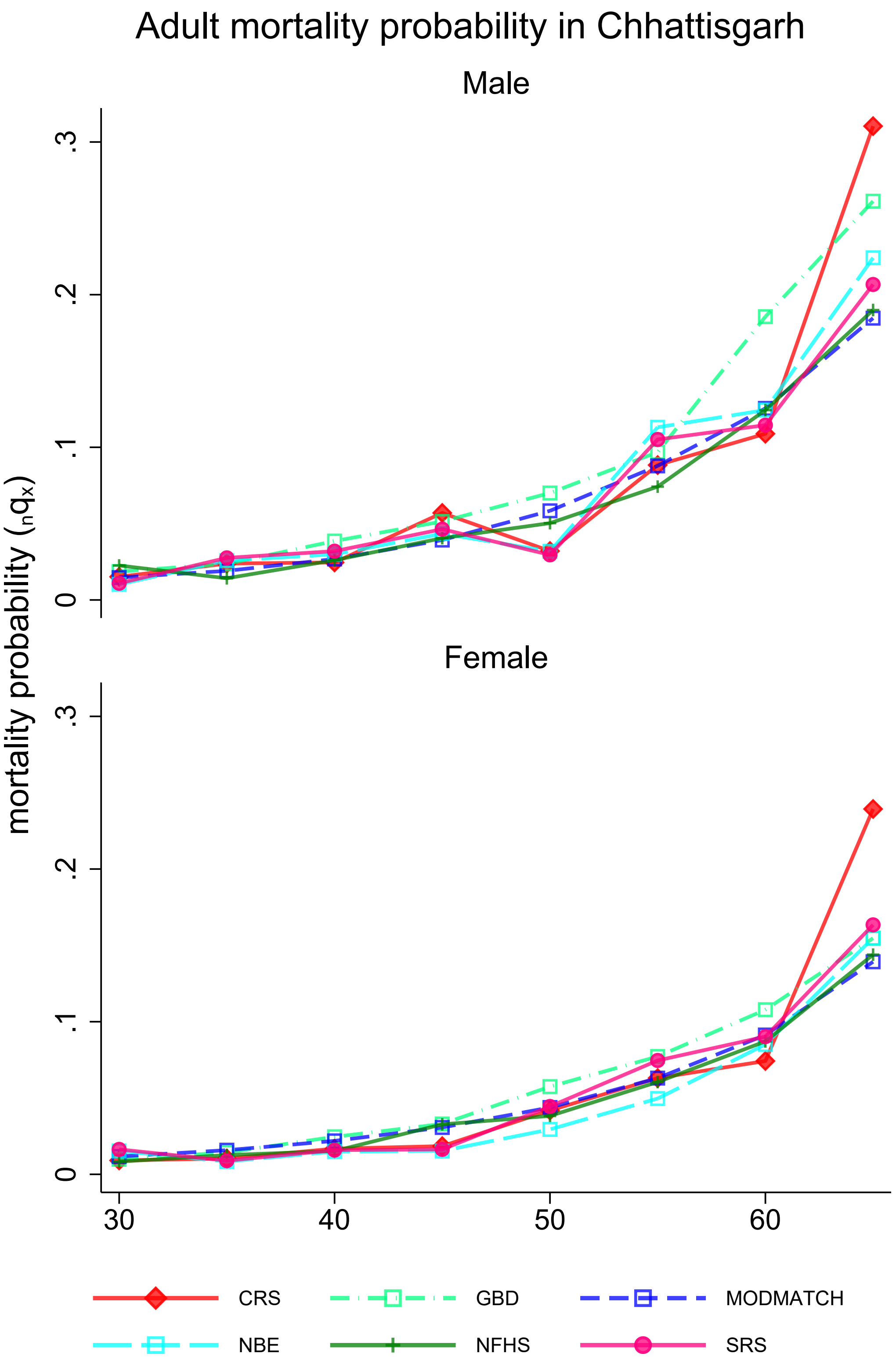

**Appendix 3E:** Mortality probabilities by place, source, and sex

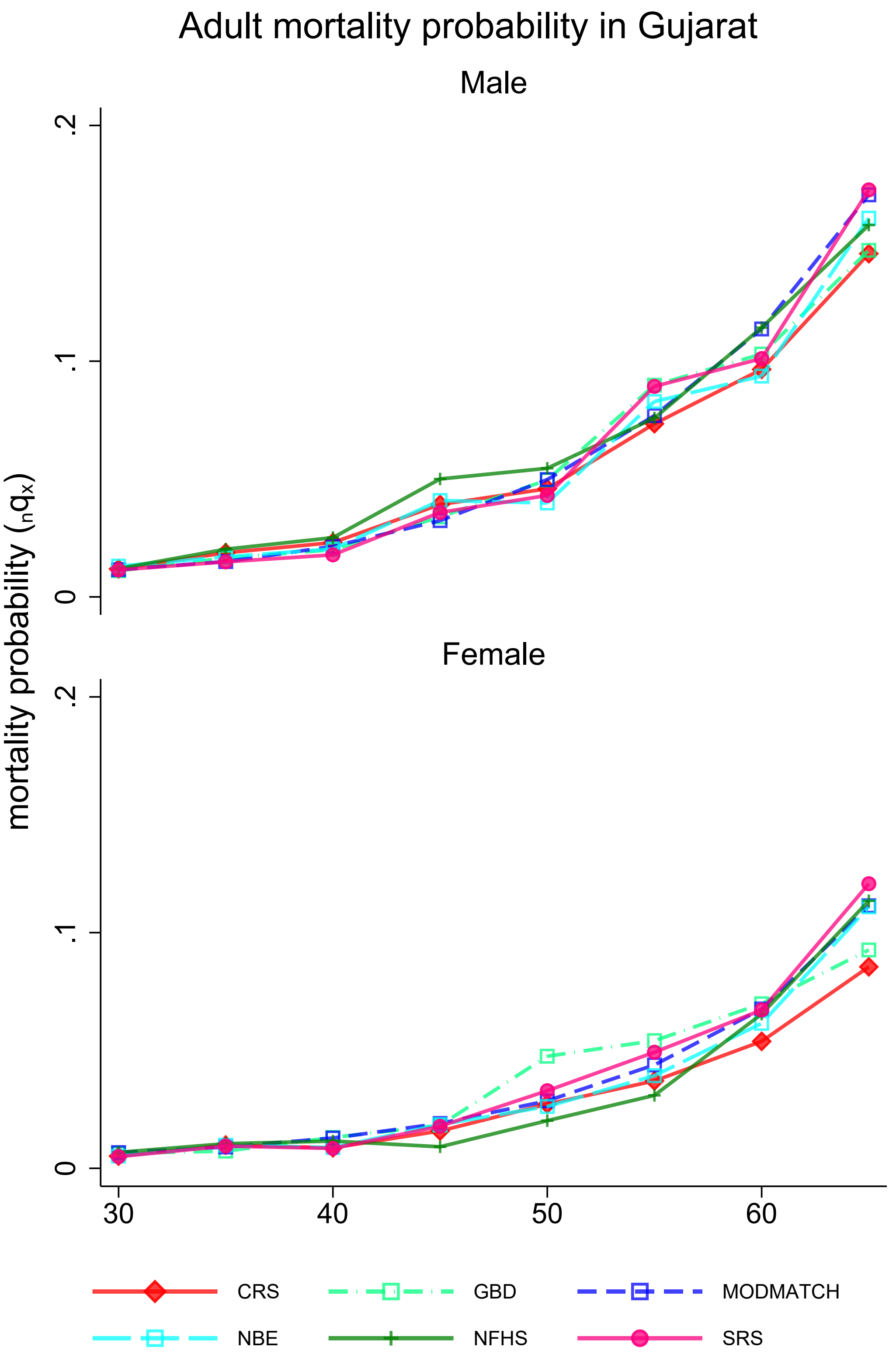

**Appendix 3F:** Mortality probabilities by place, source, and sex

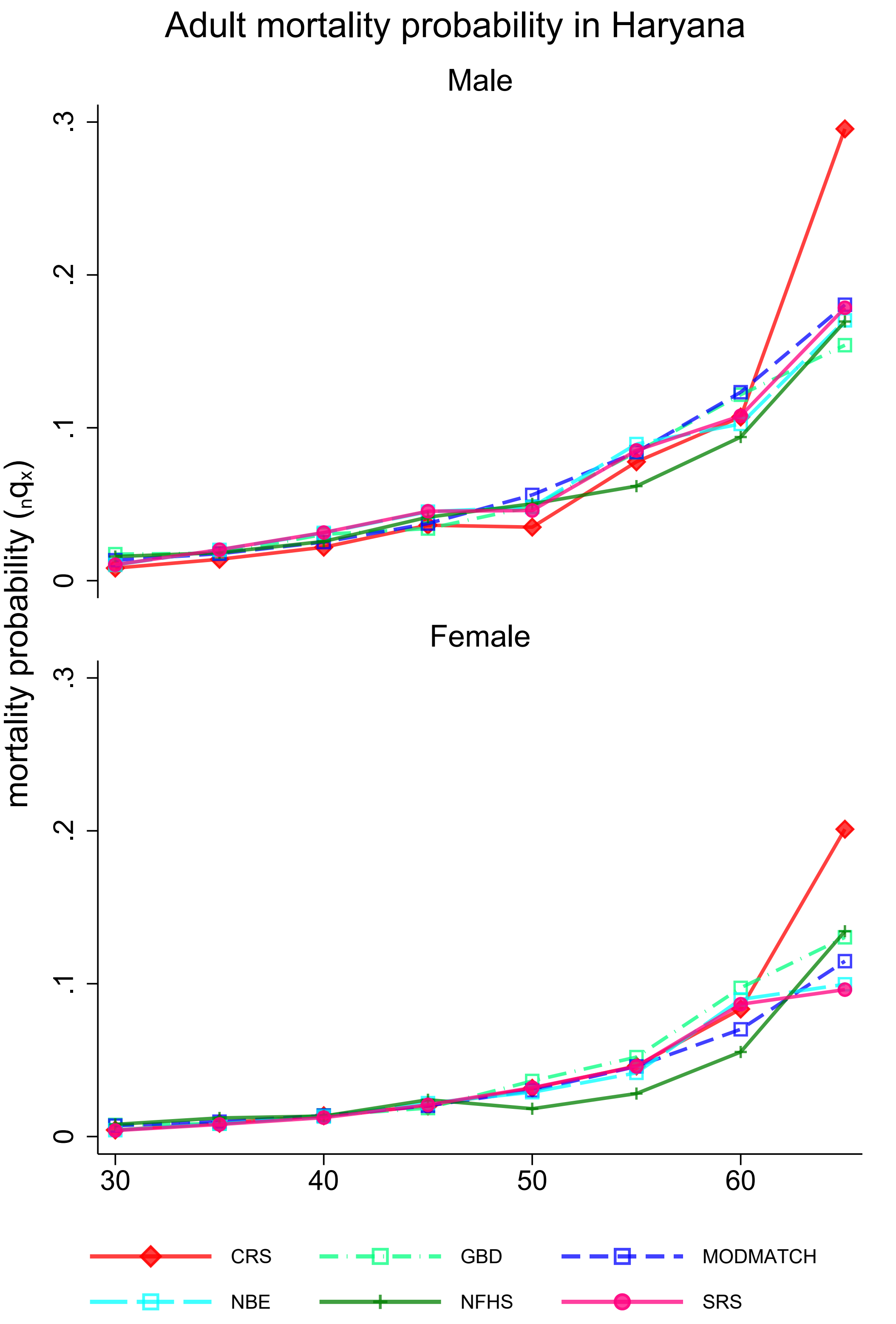

**Appendix 3G:** Mortality rates by place, source, and sex

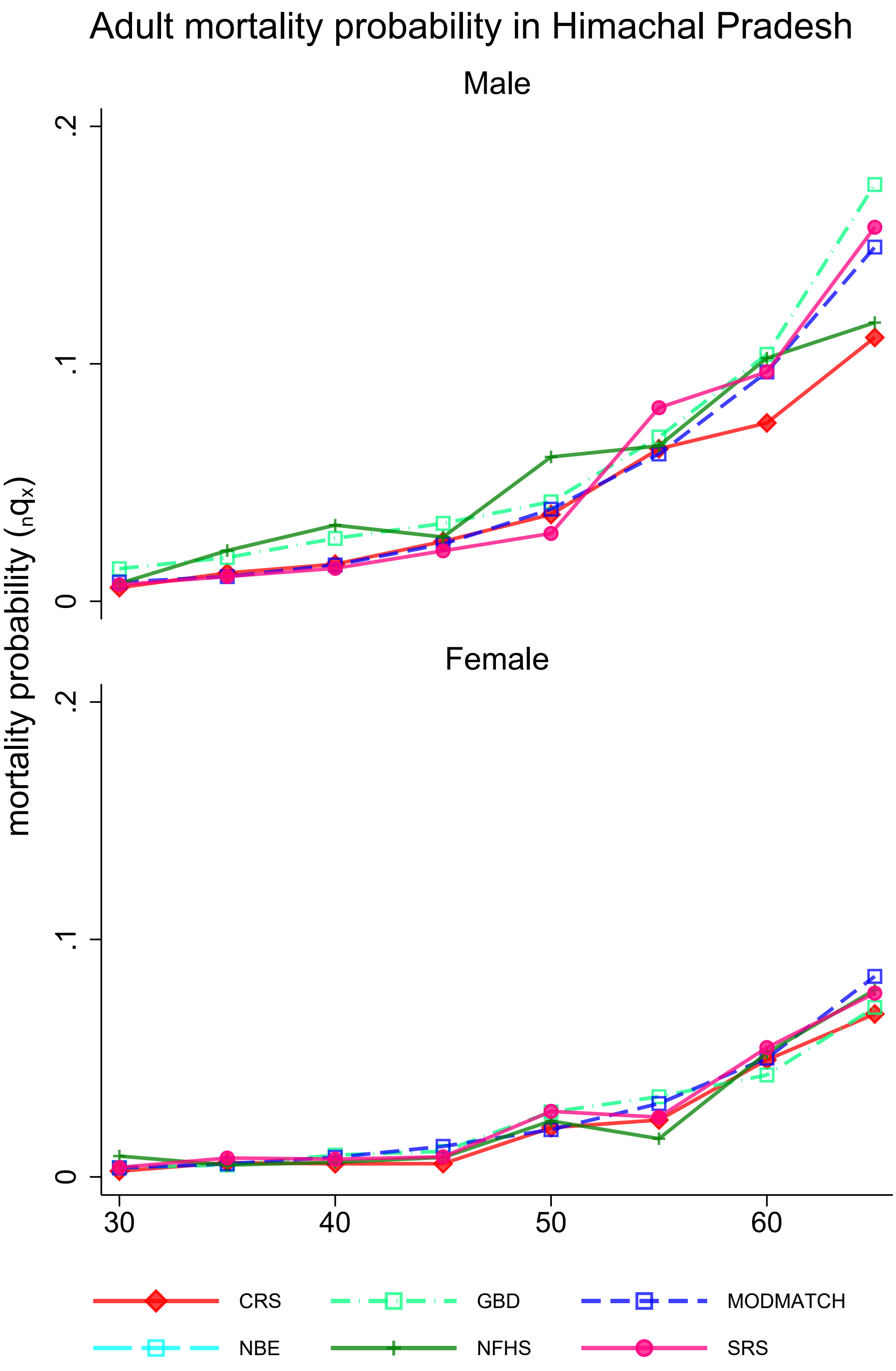

**Appendix 3H:** Mortality probabilities by place, source, and sex

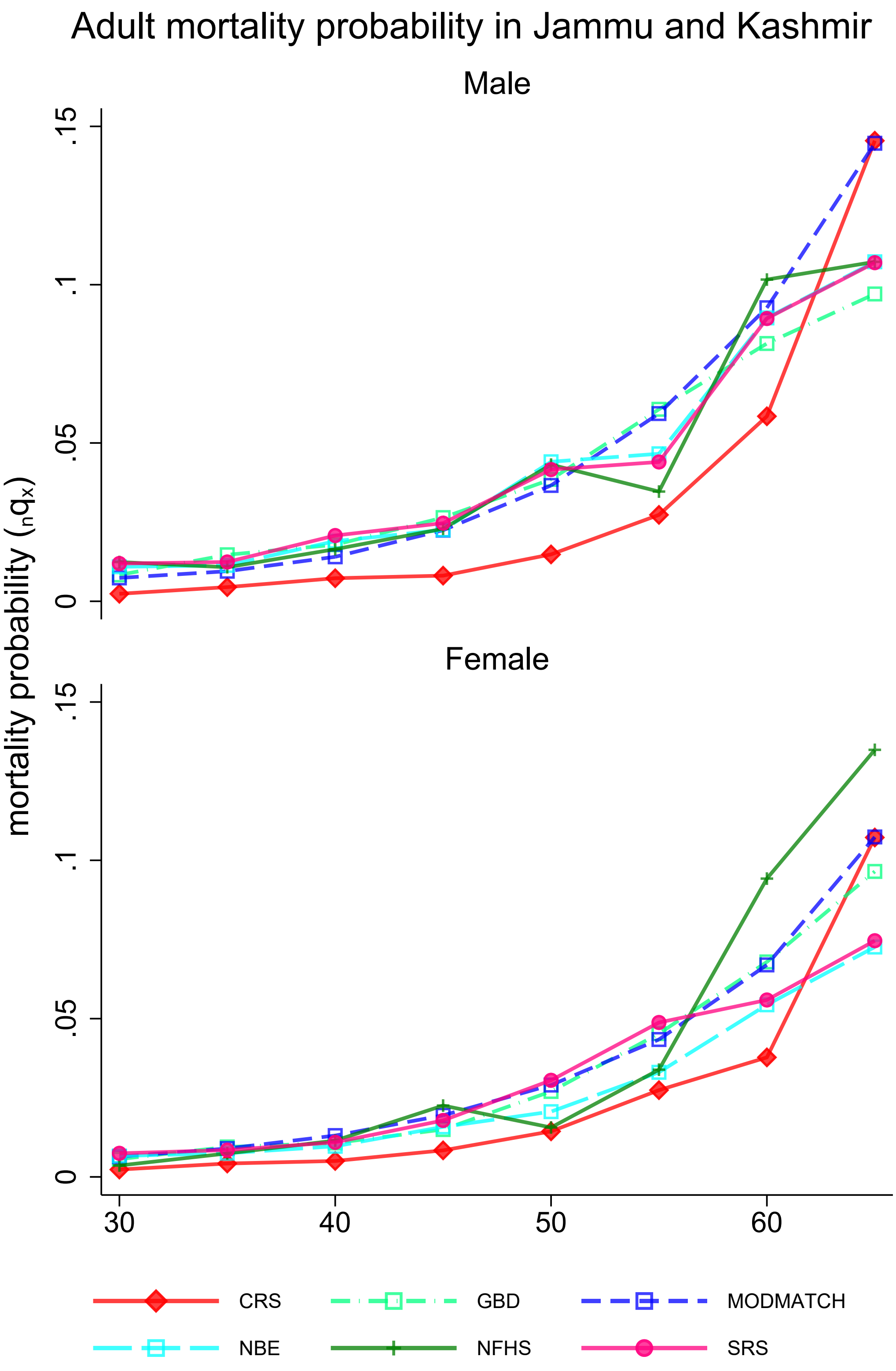

**Appendix 3I:** Mortality rates by place, source, and sex

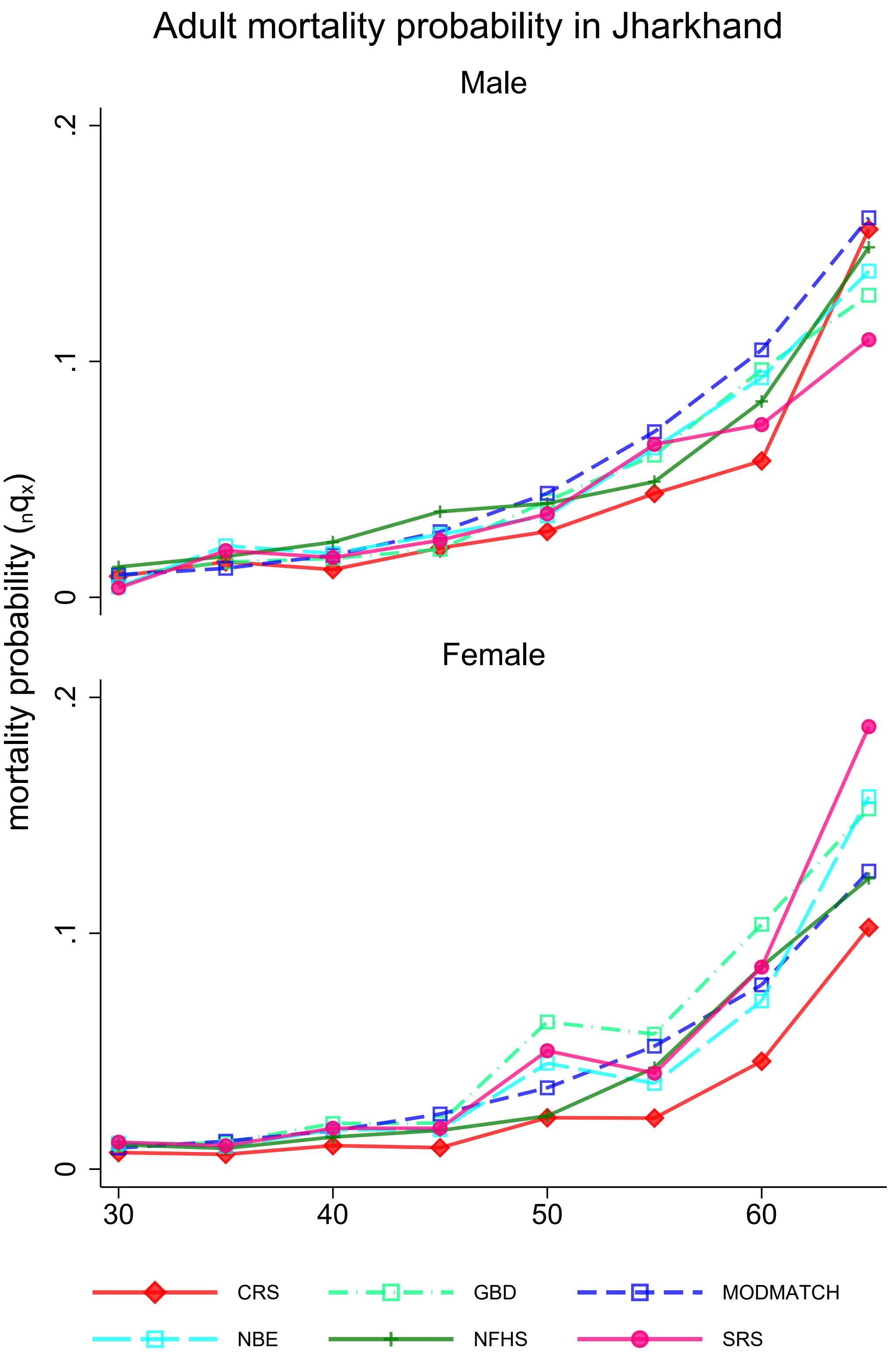

**Appendix 3J:** Mortality probabilities by place, source, and sex

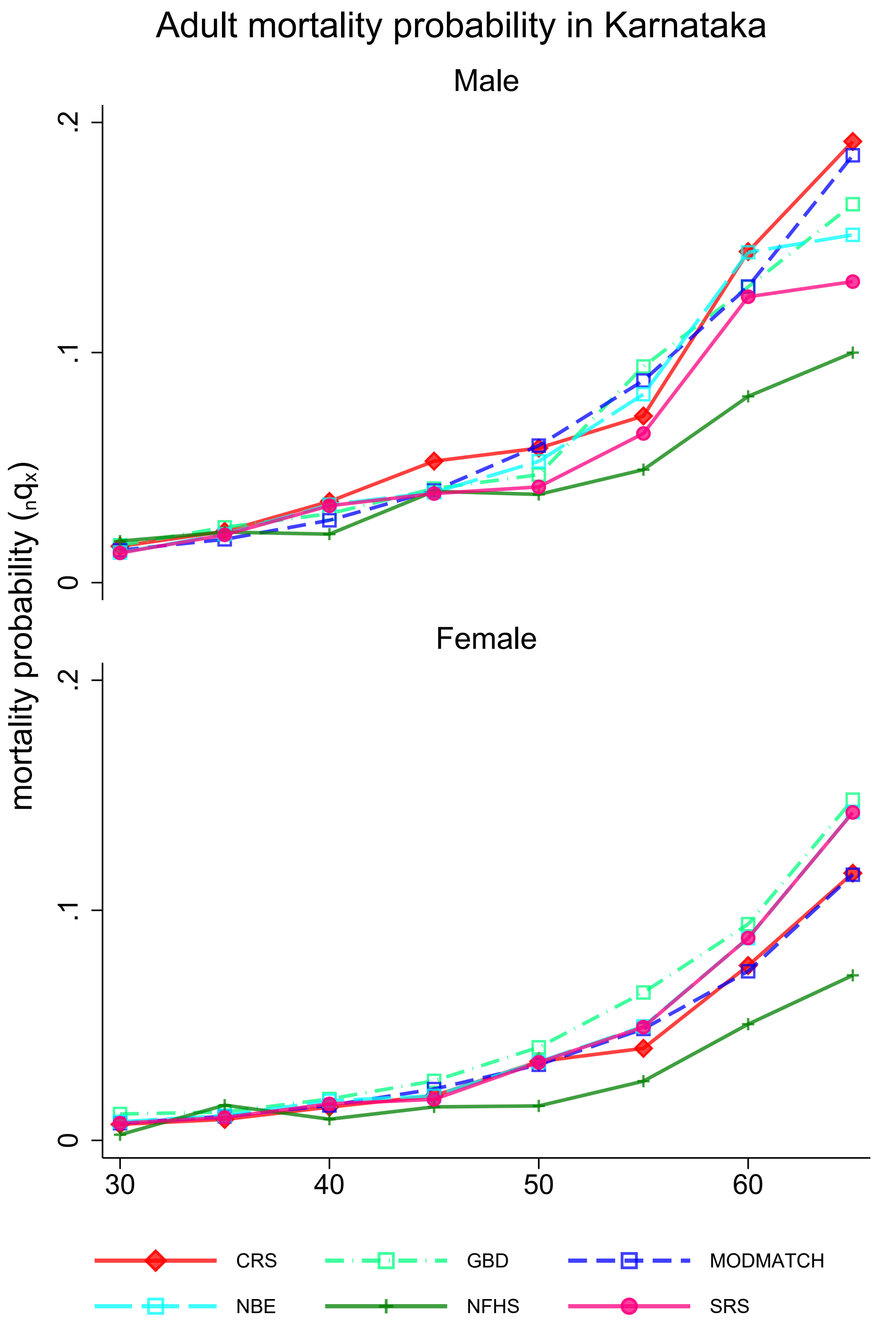

**Appendix 3K:** Mortality rates by place, source, and sex

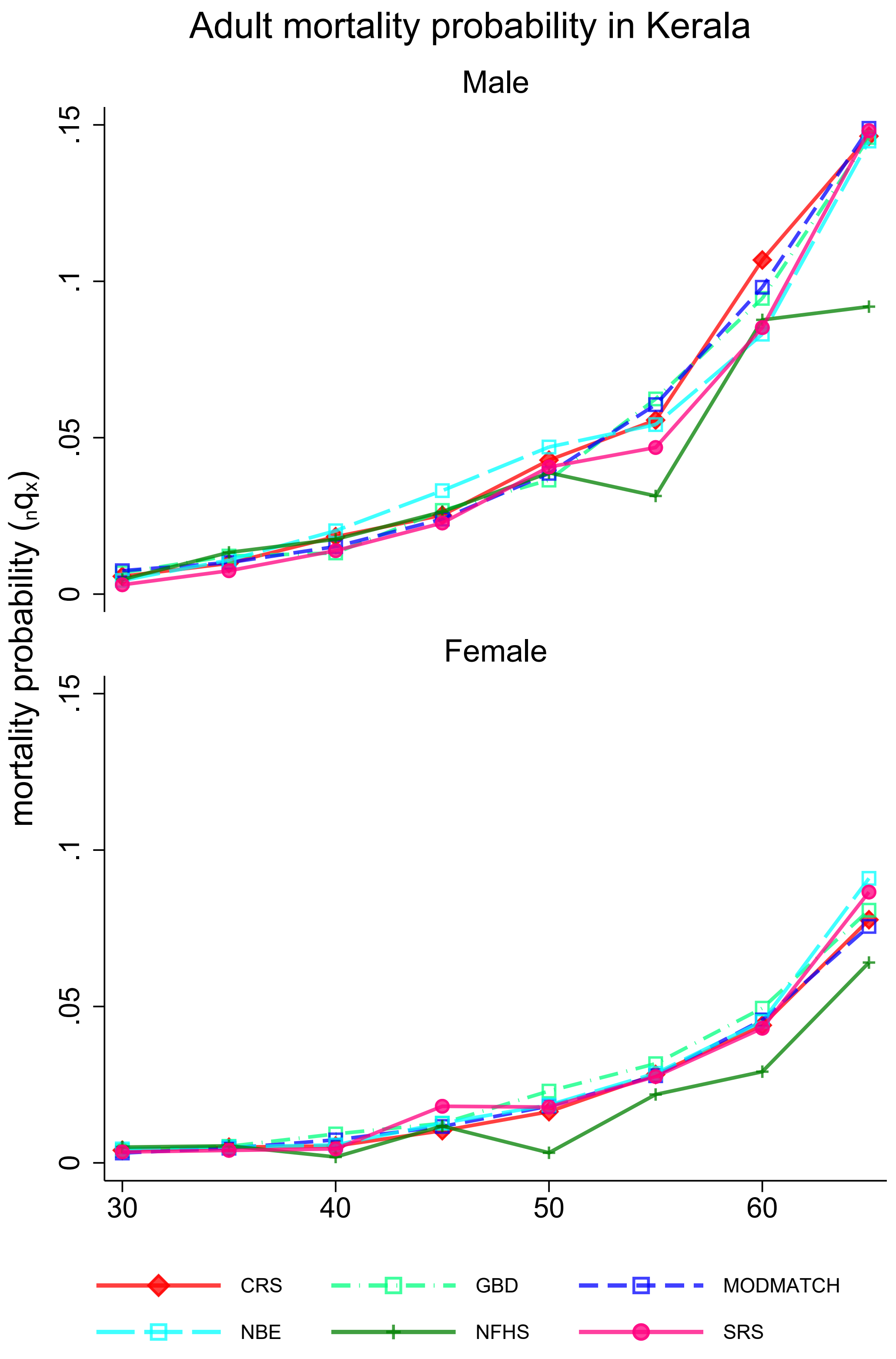

**Appendix 3L:** Mortality probabilities by place, source, and sex

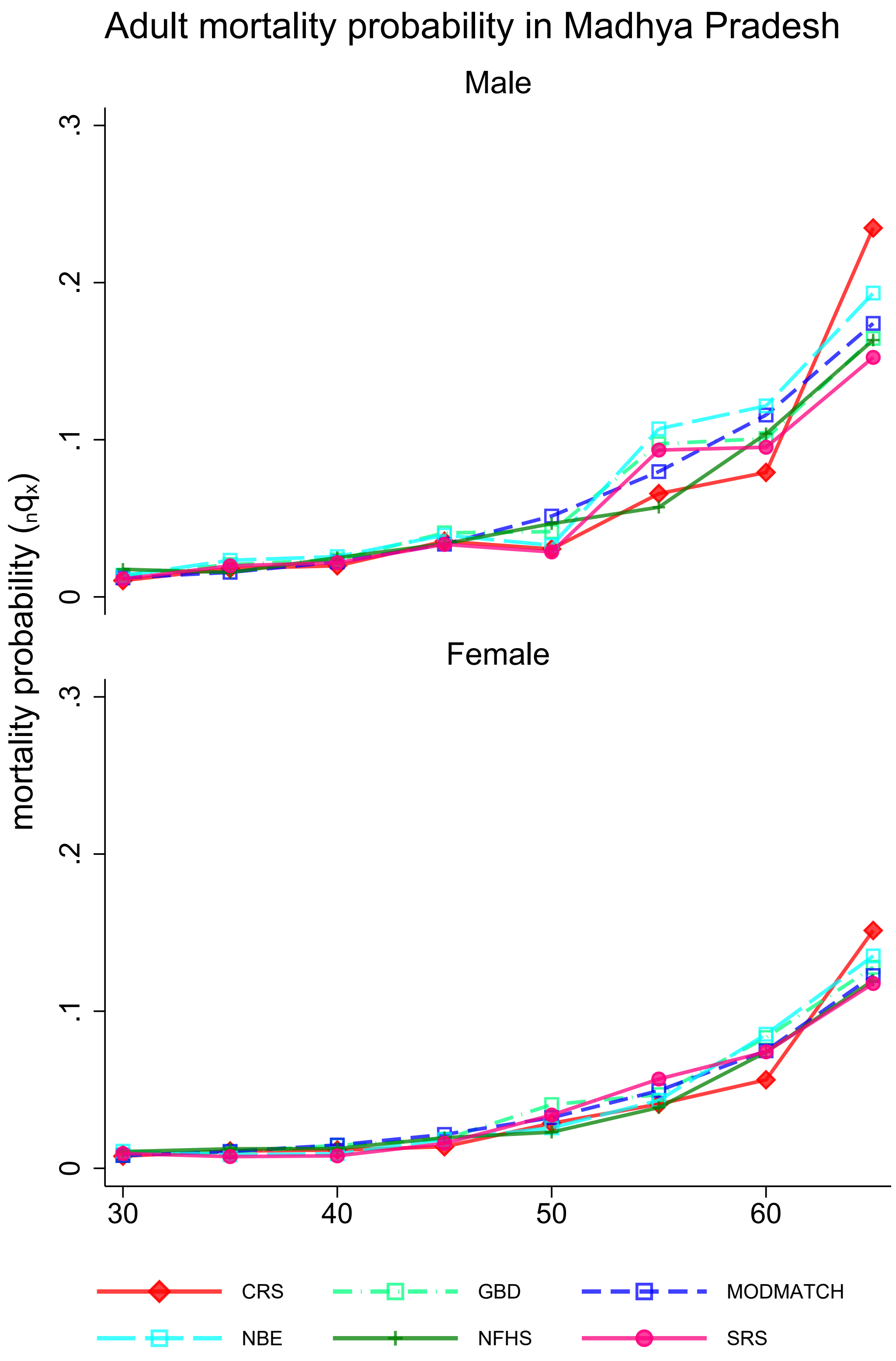

**Appendix 3M:** Mortality rates by place, source, and sex

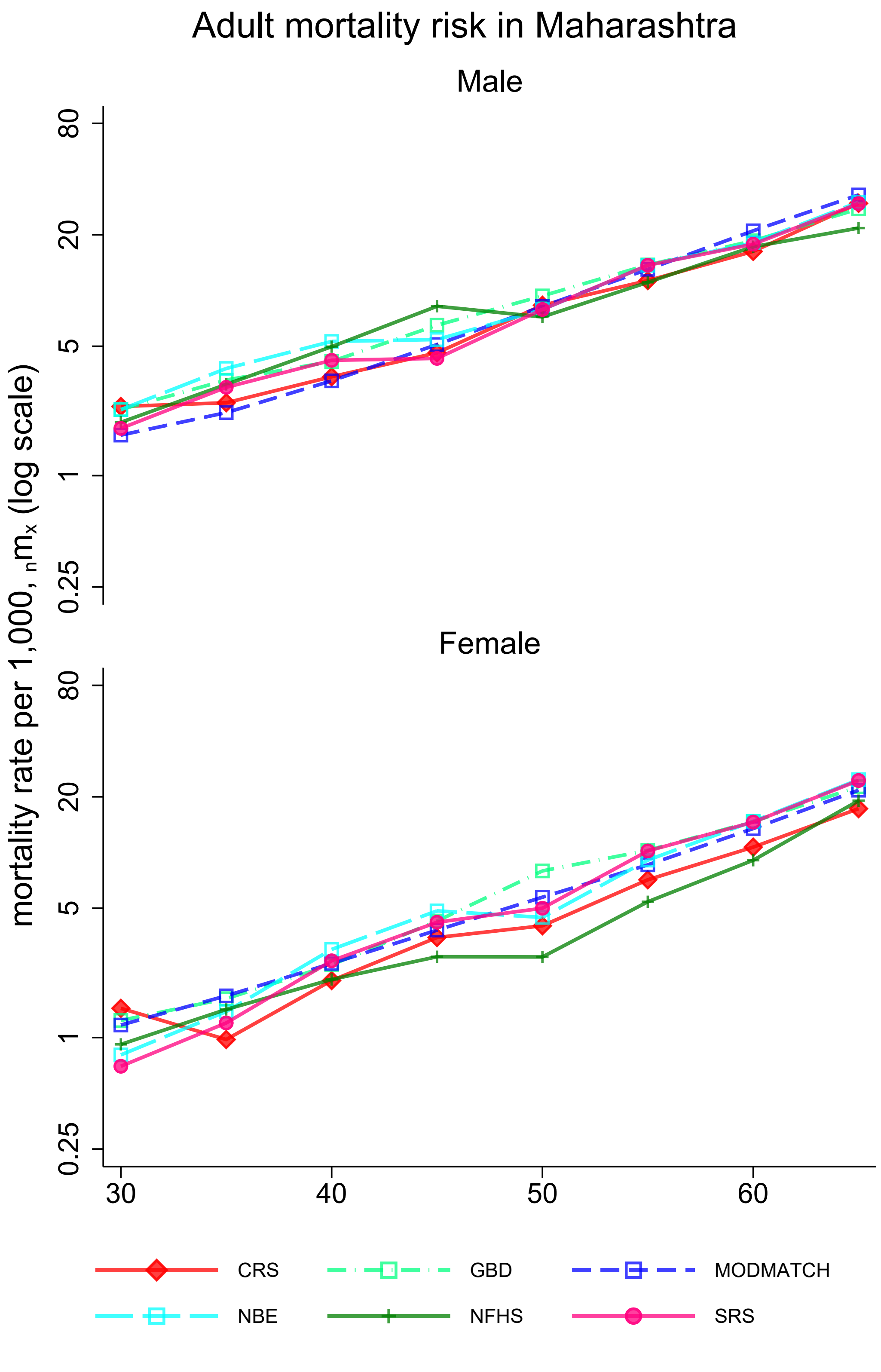

**Appendix 3N:** Mortality probabilities by place, source, and sex

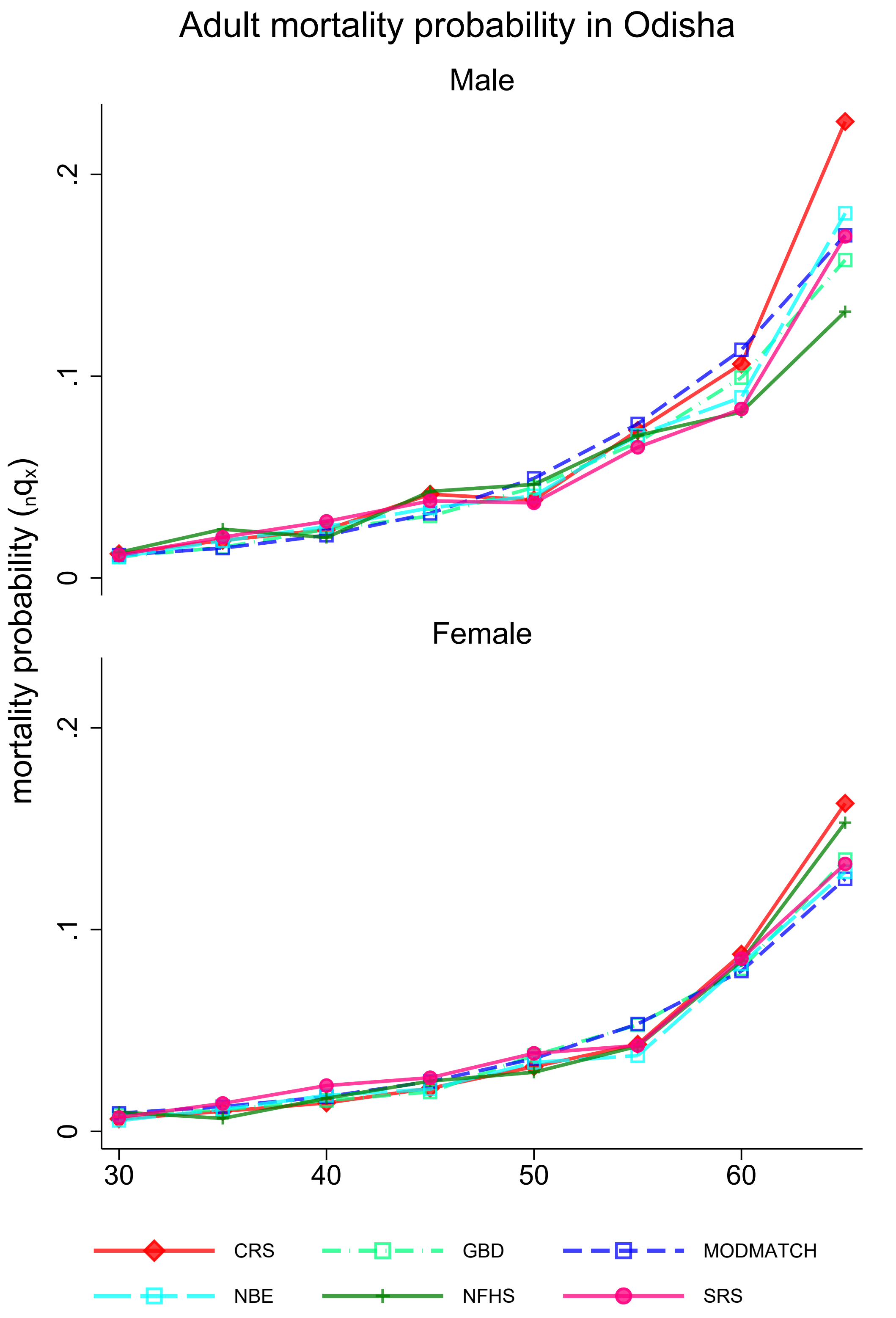

**Appendix 3O:** Mortality probabilities by place, source, and sex

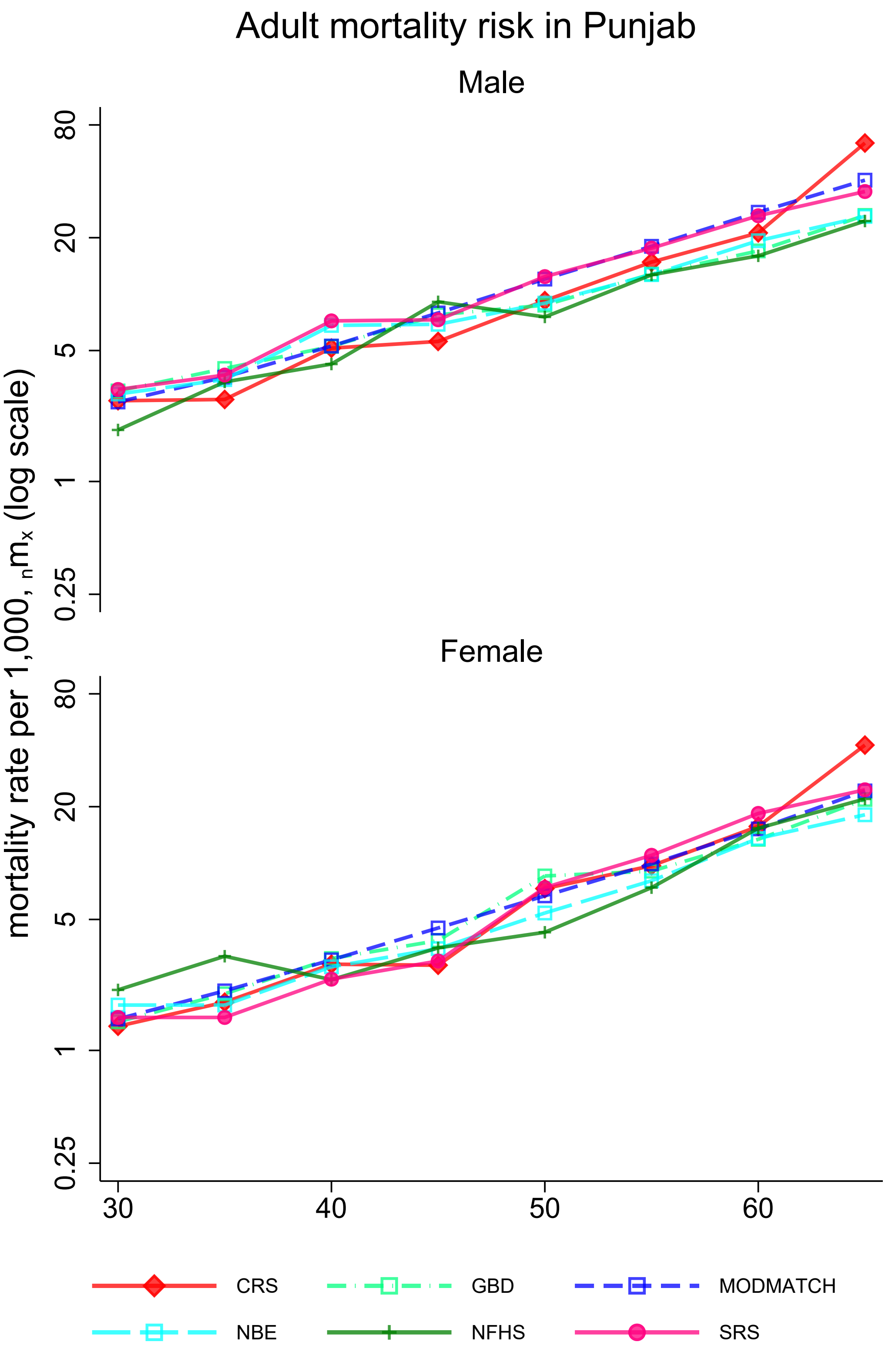

**Appendix 3P:** Mortality probabilities by place, source, and sex

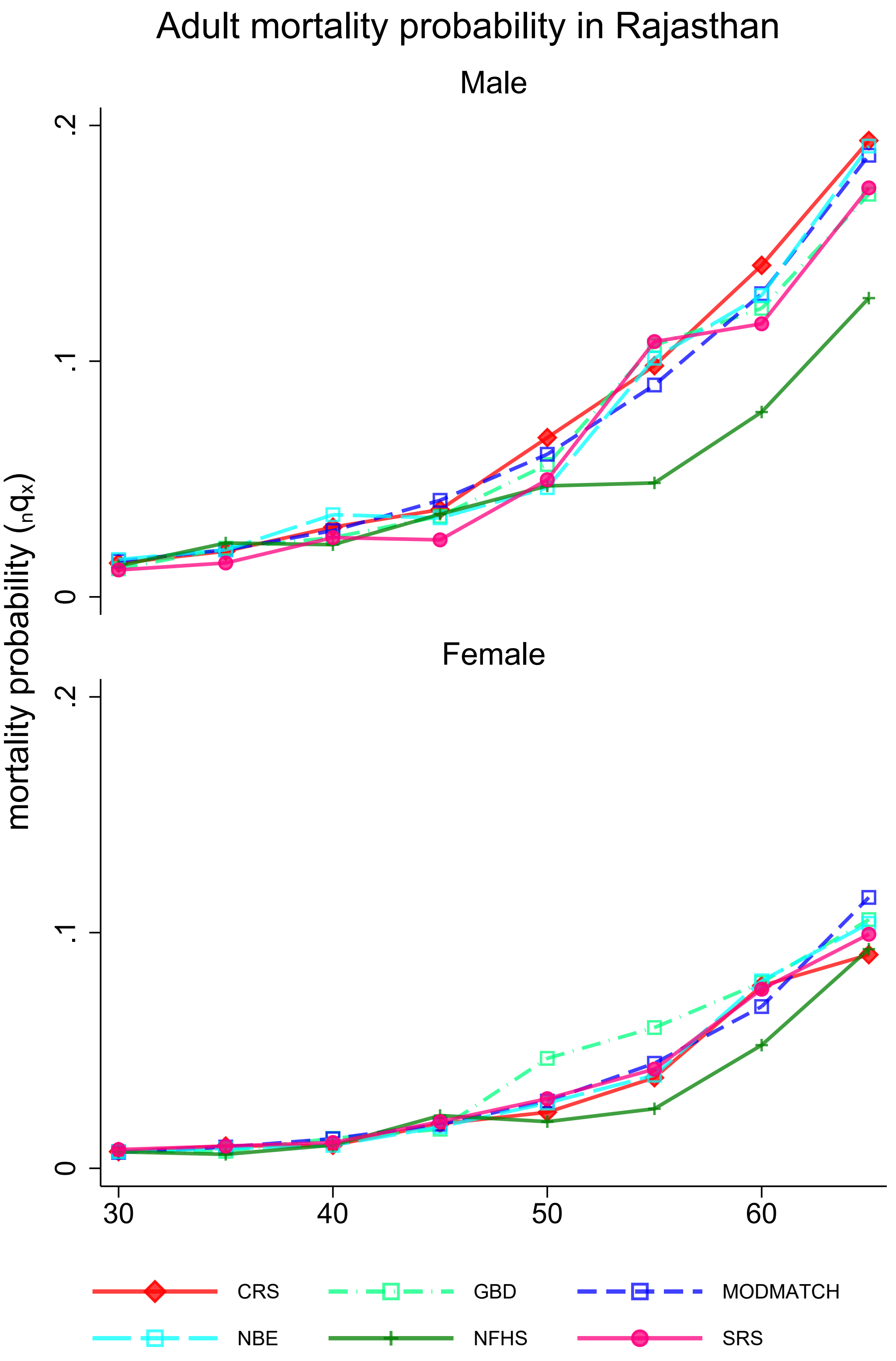

**Appendix 3Q:** Mortality probabilities by place, source, and sex

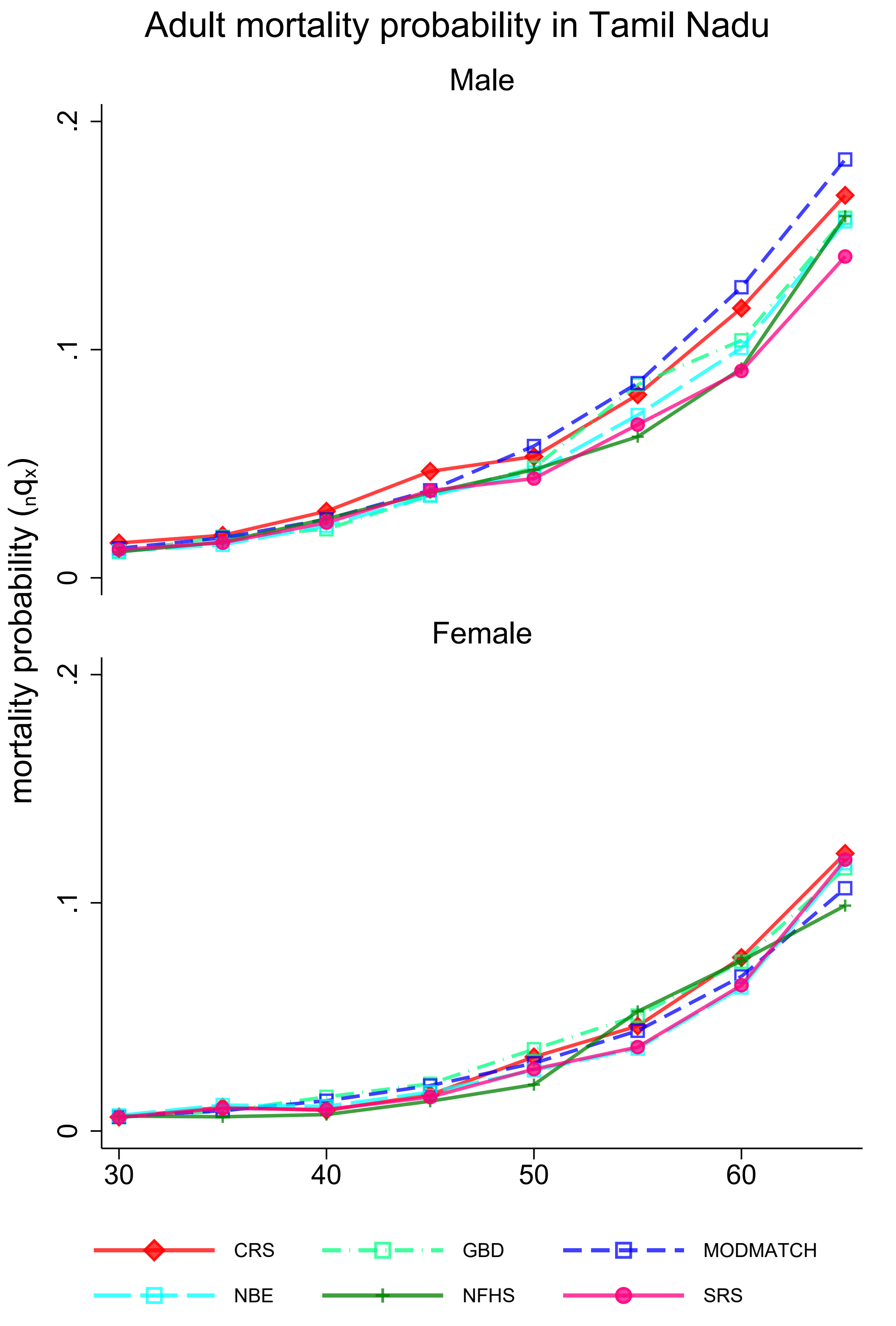

**Appendix 3R:** Mortality probabilities by place, source, and sex

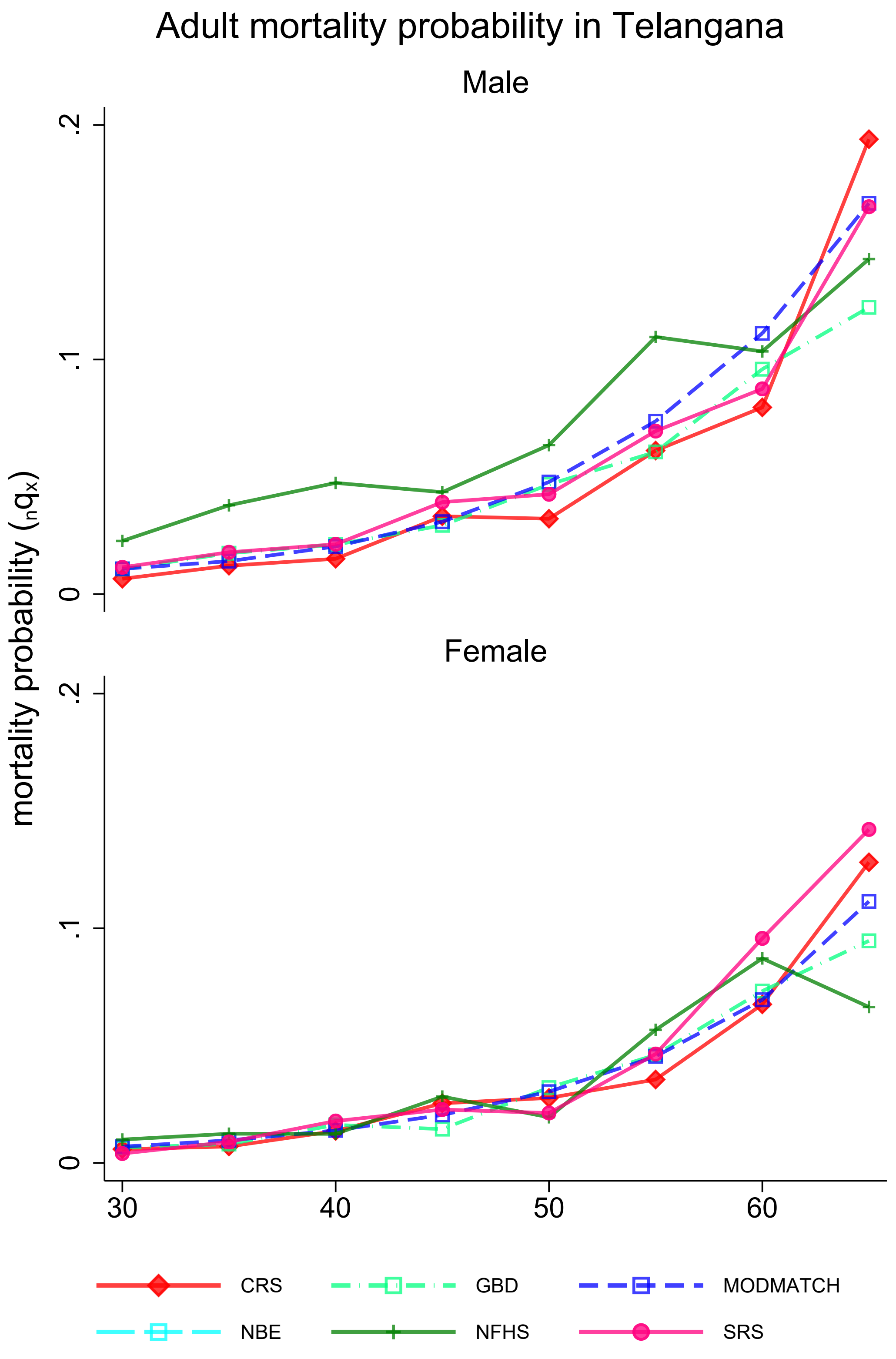

**Appendix 3S:** Mortality probabilities by place, source, and sex

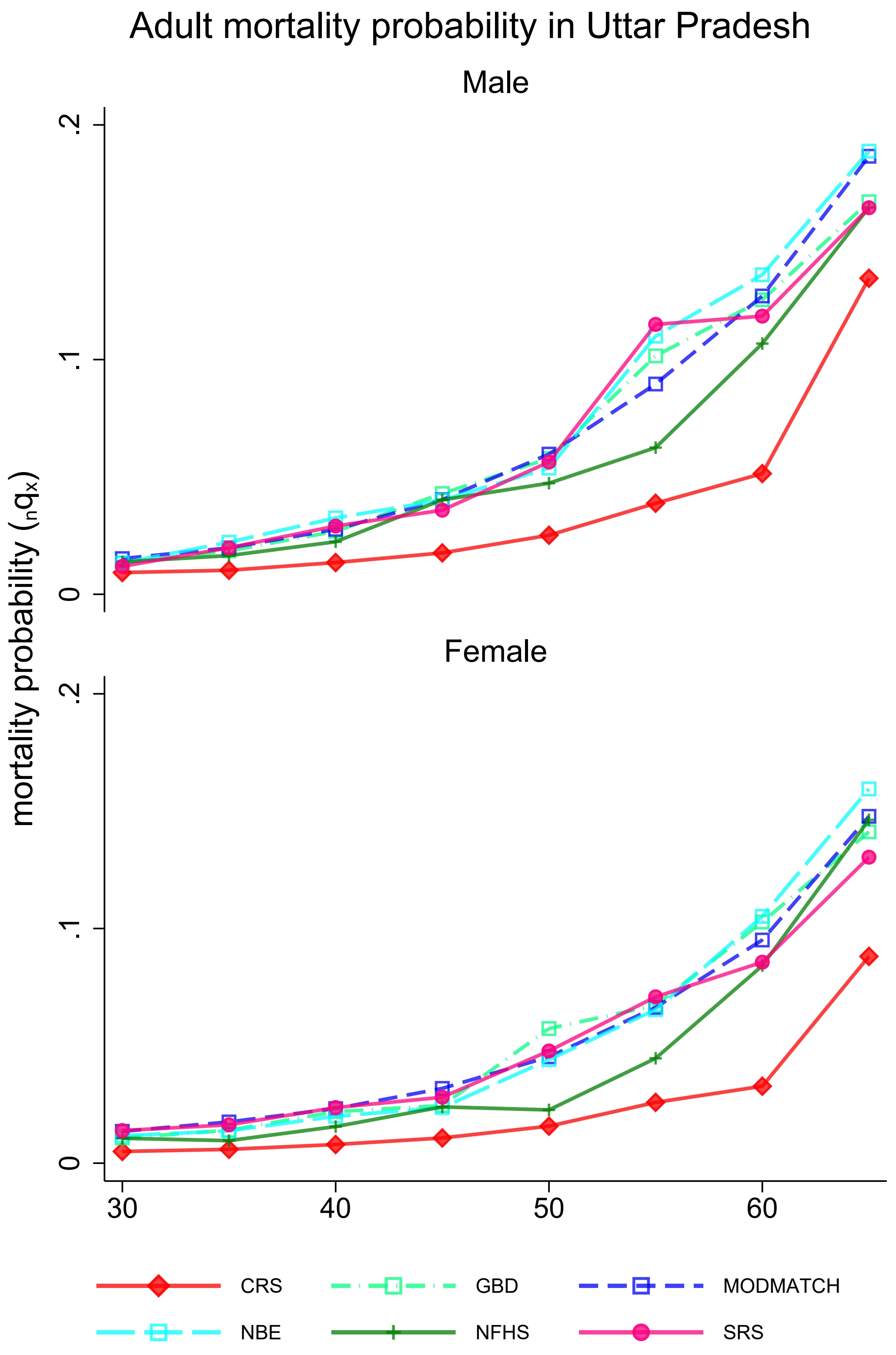

**Appendix 3T:** Mortality probabilities by place, source, and sex

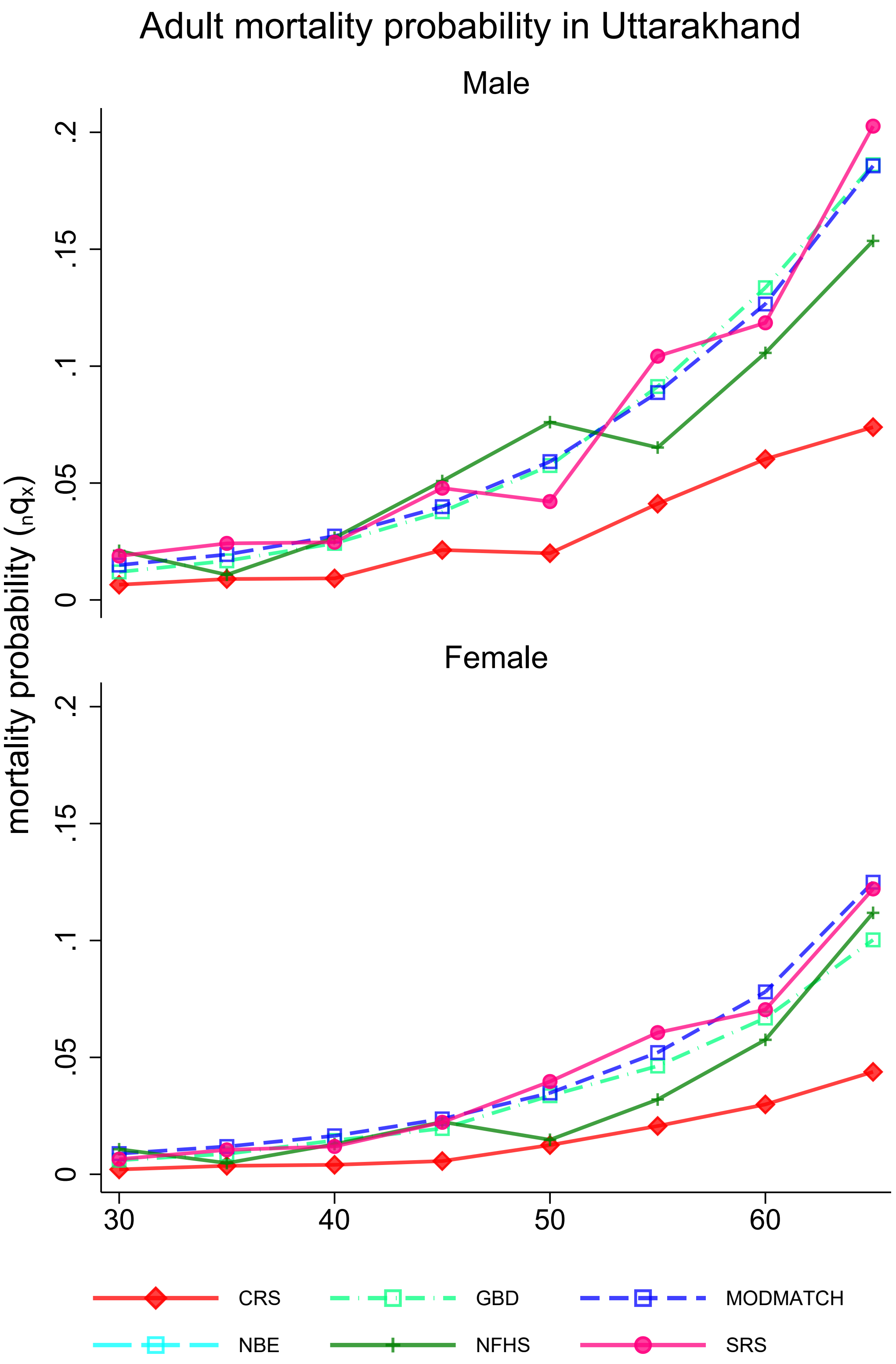

**Appendix 3U:** Mortality probabilities by place, source, and sex

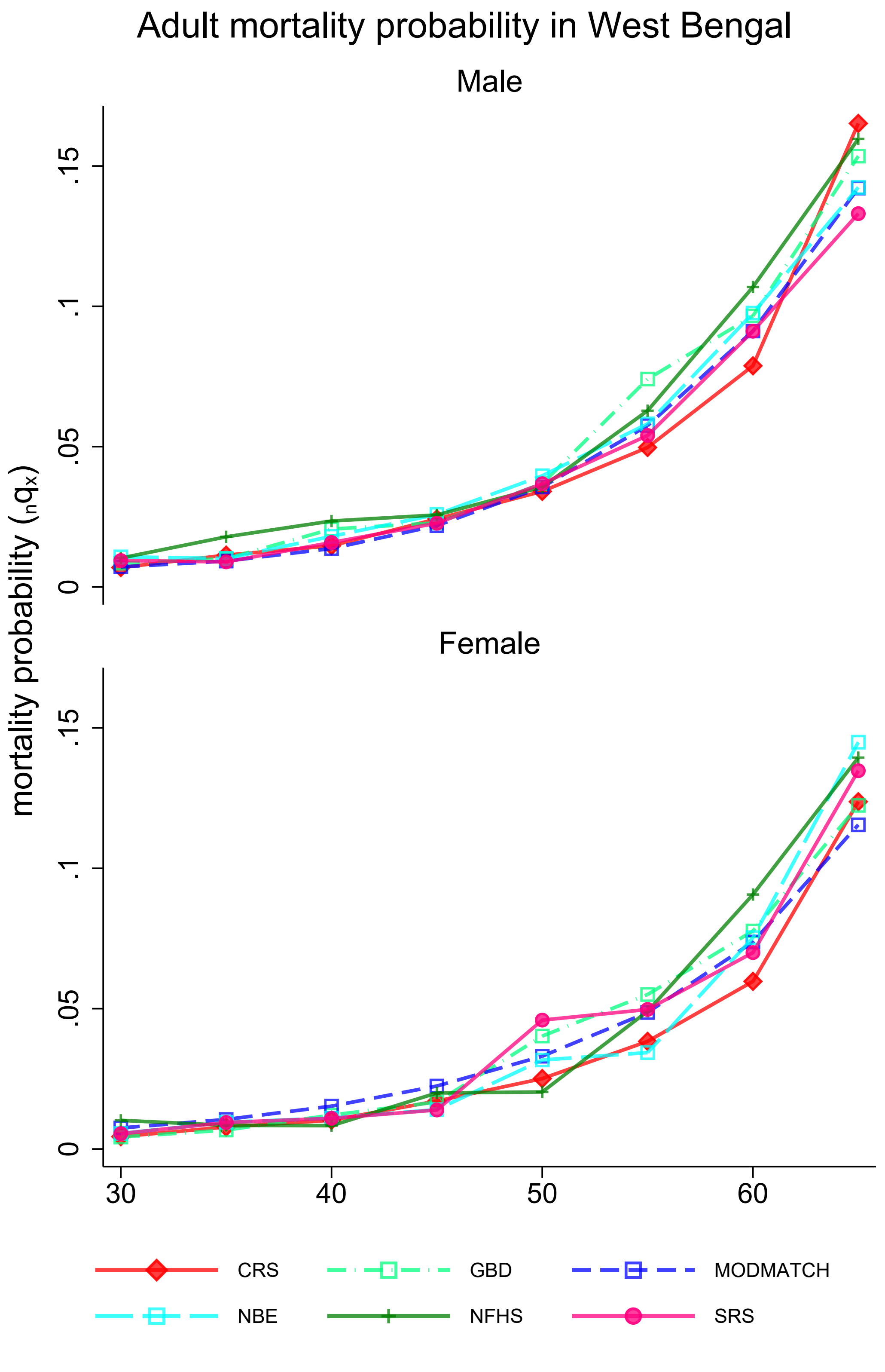

**Appendix 3V:** Mortality probabilities by place, source, and sex

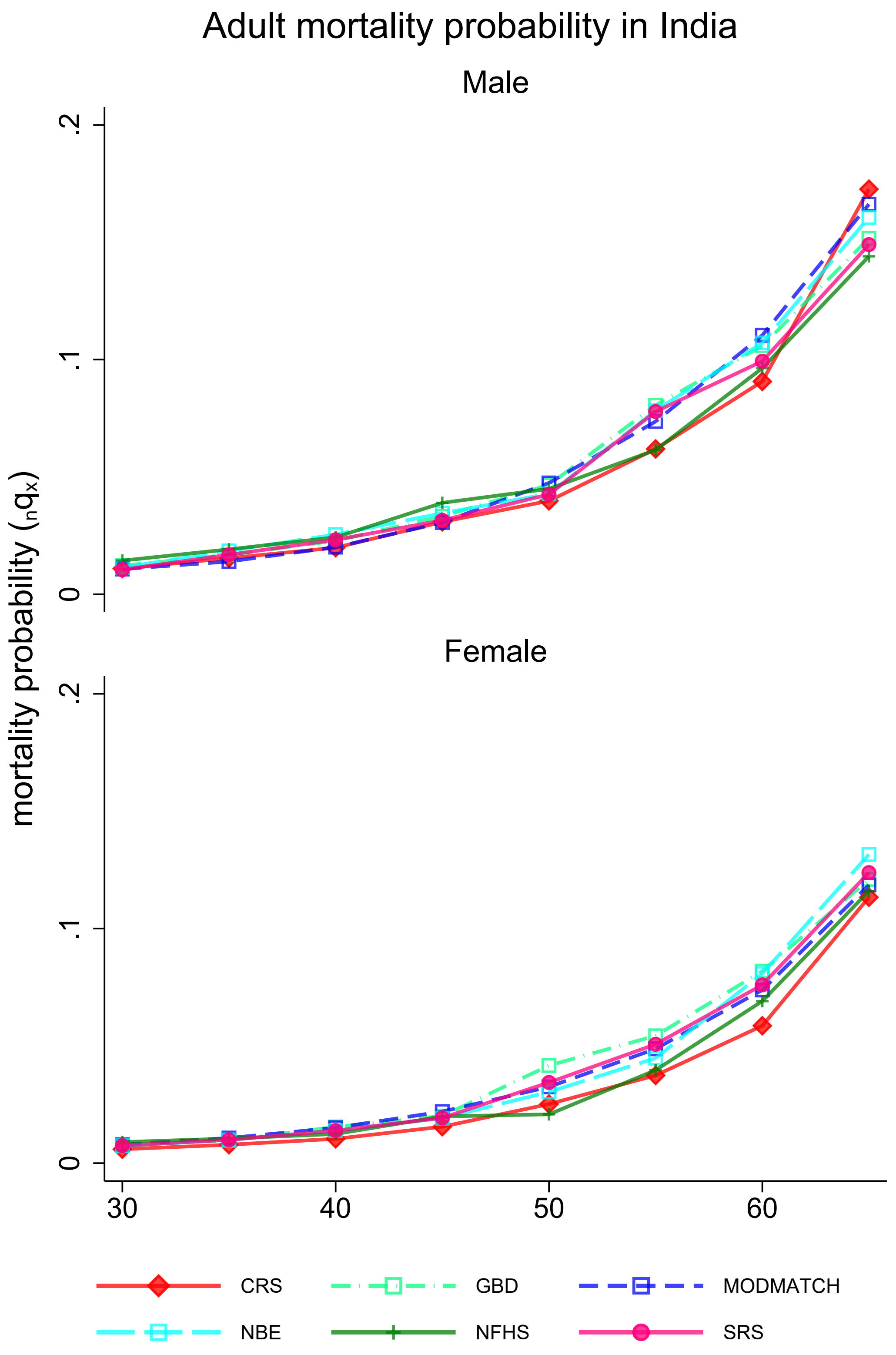
